# Impacts of vaccination and Severe Acute Respiratory Syndrome Coronavirus 2 variants Alpha and Delta on Coronavirus Disease 2019 transmission dynamics in four metropolitan areas of the United States

**DOI:** 10.1101/2021.10.19.21265223

**Authors:** Abhishek Mallela, Ye Chen, Yen Ting Lin, Ely F. Miller, Jacob Neumann, Zhili He, Kathryn E. Nelson, Richard G. Posner, William S. Hlavacek

## Abstract

To characterize Coronavirus Disease 2019 (COVID-19) transmission dynamics in each of the metropolitan statistical areas (MSAs) surrounding Dallas, Houston, New York City, and Phoenix in 2020 and 2021, we extended a previously reported compartmental model accounting for effects of multiple distinct periods of non-pharmaceutical interventions by adding consideration of vaccination and Severe Acute Respiratory Syndrome Coronavirus 2 (SARS-CoV-2) variants Alpha (lineage B.1.1.7) and Delta (lineage B.1.617.2). For each MSA, we found region-specific parameterizations of the model using daily reports of new COVID-19 cases available from January 21, 2020 to October 31, 2021. In the process, we obtained estimates of the relative infectiousness of Alpha and Delta as well as their takeoff times in each MSA (the times at which sustained transmission began). The estimated infectiousness of Alpha ranged from 1.1x to 1.4x that of viral strains circulating in 2020 and early 2021. The estimated relative infectiousness of Delta was higher in all cases, ranging from 1.6x to 2.1x. The estimated Alpha takeoff times ranged from February 1 to February 28, 2021. The estimated Delta takeoff times ranged from June 2 to June 26, 2021. Estimated takeoff times are consistent with genomic surveillance data.

**One-Sentence Summary:** Using a compartmental model parameterized to reproduce available reports of new Coronavirus Disease 2019 (COVID-19) cases, we quantified the impacts of vaccination and Severe Acute Respiratory Syndrome Coronavirus 2 (SARS-CoV-2) variants Alpha (lineage B.1.1.7) and Delta (lineage B.1.617.2) on regional epidemics in the metropolitan statistical areas (MSAs) surrounding Dallas, Houston, New York City, and Phoenix.

## INTRODUCTION

In 2020, Coronavirus Disease 2019 (COVID-19) transmission dynamics were significantly influenced by non-pharmaceutical interventions [1–7]. In 2021, other factors arose with significant impacts on disease transmission, namely, vaccination [8–9] and emergence of Severe Acute Respiratory Syndrome Coronavirus 2 (SARS-CoV-2) variants [10–11].

Mass vaccination in the United States (US) began on December 14, 2020 [12], with demonstrable reduction of disease burden within vaccinated populations [13]. As the vaccination campaign progressed into March 2021, there was widespread reduction in disease incidence [14] and relaxation of state-mandated non-pharmaceutical interventions [15].

In early 2021, SARS-CoV-2 variant Alpha (lineage B.1.1.7) spread across the US and became the dominant circulating strain [16]. By the end of July 2021, the Delta variant (lineage B.1.617.2) had supplanted Alpha [17], concomitant with increases in new COVID-19 case detection [14]. Both Alpha and Delta have been estimated to be more transmissible than strains circulating earlier [18–23], and it was determined that vaccinated persons infected with Alpha and Delta were capable of transmitting disease [24, 25].

The Alpha variant was first detected in Kent, England in September 2020 [26], and was declared a variant of concern (VOC) on December 18, 2020 [26]. The Delta variant was first detected in Maharashtra, India in October 2020 [27] and was declared a VOC on May 6, 2021 [26]. In the literature, previous modeling works have investigated the relative transmissibility and timing of SARS-CoV-2 variants Alpha, Delta, and Omicron in multiple regions/countries, including England, Greece, Iran, China, and the US [28–36]. Modeling approaches applied in these studies involved deterministic [32–34] and stochastic [28, 30] compartmental models of COVID-19 transmission. Other approaches involved statistical models and various forms of regression, e.g., multinomial logistic regression [29] and multivariable binary hyperbolastic regression [36]. Bayesian methods, including the sequential Bayesian method [32], a Bayesian evidence synthesis framework [30], a Bayesian approach to estimate the effective reproduction number *R*_*t*_ [31] and Markov Chain Monte Carlo (MCMC) sampling [28, 33], were predominantly used. The data considered in these studies included case data [28, 30–35] viral load data [28], seroprevalence data [30], information on deaths and hospital admissions [33], age-specific vaccine coverage data [31], sequencing data [29], and biospecimen data [35].

In earlier work, we demonstrated that new COVID-19 case detection over various periods in 2020 can be faithfully reproduced for 280 (out of the 384) metropolitan statistical areas (MSAs) in the US and all 50 states by region-specific parameterizations of a compartmental model that accounts for time-varying non-pharmaceutical interventions [5–7]. We found that the multiple surges in disease incidence seen in 2020 [14] could be explained by changes in protective disease-avoiding behaviors, which we will refer to collectively as social-distancing.

However, in 2021, the model lost its ability to capture disease transmission dynamics, presumably because of the impacts of vaccination and the emergence of more transmissible SARS-CoV-2 variants, namely, Alpha and Delta. Here, to quantify the impacts of vaccination and SARS-CoV-2 variants Alpha and Delta on COVID-19 transmission dynamics, we extended the model of Lin et al. [5] by adding consideration of vaccination and variants with increased transmissibility. We then found region-specific parameterizations of the model using vaccination and surveillance case data available for the MSAs surrounding Dallas, Houston, New York City, and Phoenix.

## METHODS

### Data

Daily reports of new confirmed COVID-19 cases were obtained from the GitHub repository maintained by *The New York Times* newspaper [37]. Daily reports of newly completed vaccinations were obtained from the Covid Act Now database for the MSAs surrounding New York City and Phoenix [38]. Because of reporting gaps in the Covid Act Now database, we used a different source of vaccination data for the MSAs surrounding Dallas and Houston, the *Democrat and Chronicle* newspaper [39]. County-level surveillance and vaccination data were aggregated to obtain daily case and vaccination counts for the MSAs surrounding Dallas, Houston, New York City, and Phoenix. In the case of a missing daily report, we imputed the missing information as described in the Appendix.

### Compartmental Model for Disease Transmission Dynamics

We used the compartmental model illustrated in Figure 1 (and Appendix Figure 1) to analyze data available for each MSA of interest. The model consists of ordinary differential equations (ODEs) describing the dynamics of 40 populations (state variables) (Appendix Equations 1–38). The state variables are each defined in Appendix Table 1. Model parameters are defined in Tables 1–3. Key features of the model are described below, and a full description of the model is provided in the Appendix.

**Figure 1.**
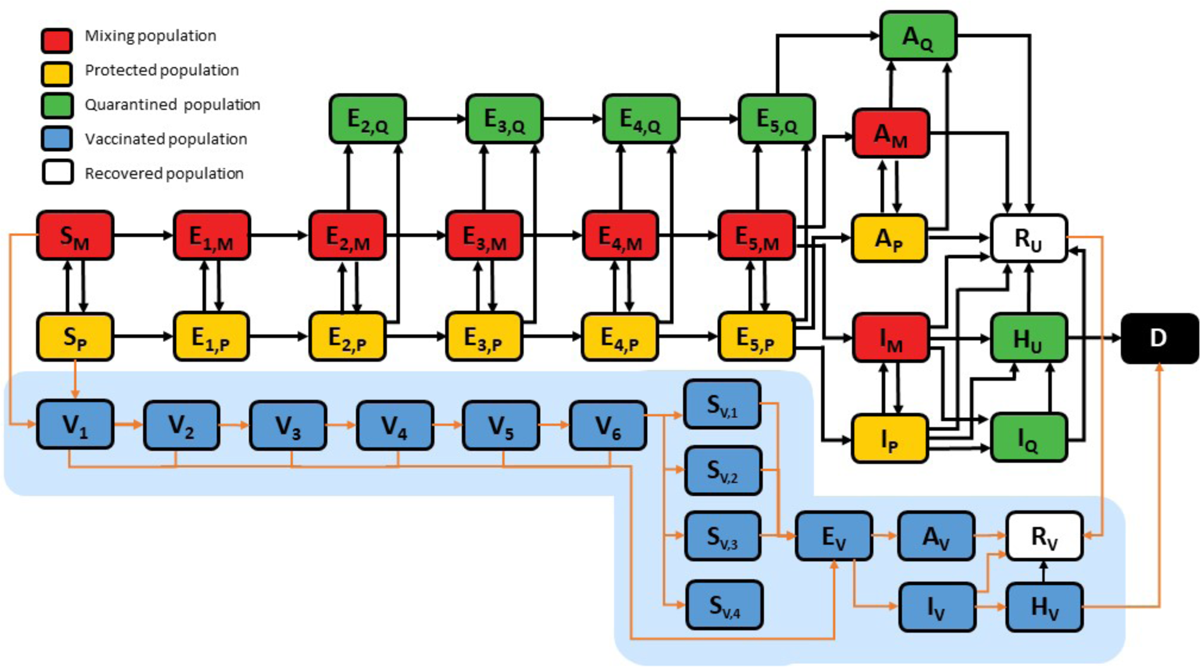
Illustration of compartmental model. The independent variable of the model is time *t*. The 40 dependent state variables of the model are populations, which are represented as boxes with rounded corners. A description of each state variable is given in Appendix Table 1. The 15 highlighted boxes (on the blue background) represent state variables introduced to capture the effects of vaccination and the Alpha and Delta variants. The other 25 boxes represent state variables considered in the model of Lin et al. [5]. Arrows connecting boxes represent transitions. Each transition represents the movement of persons from one population to another. The arrows highlighted in orange represent transitions introduced to capture the effects of vaccination and the Alpha and Delta variants. Other arrows represent transitions considered in the model of Lin et al. [5]. Each arrow is associated with one or more parameters that characterize a rate of movement; these parameters are not shown here but are shown in Appendix Figure 1. A full description of the model is given in the Appendix.

**Table 1.**
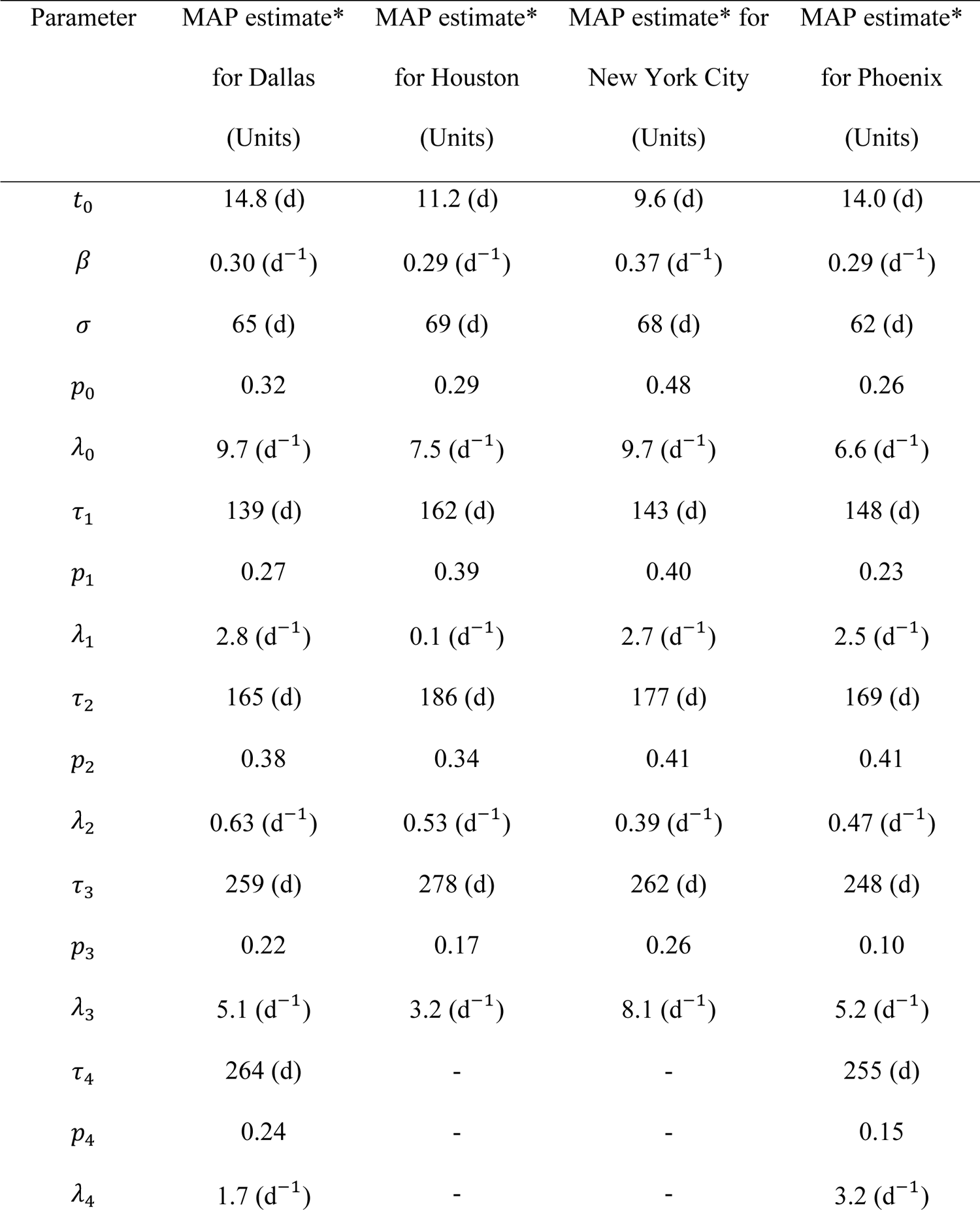
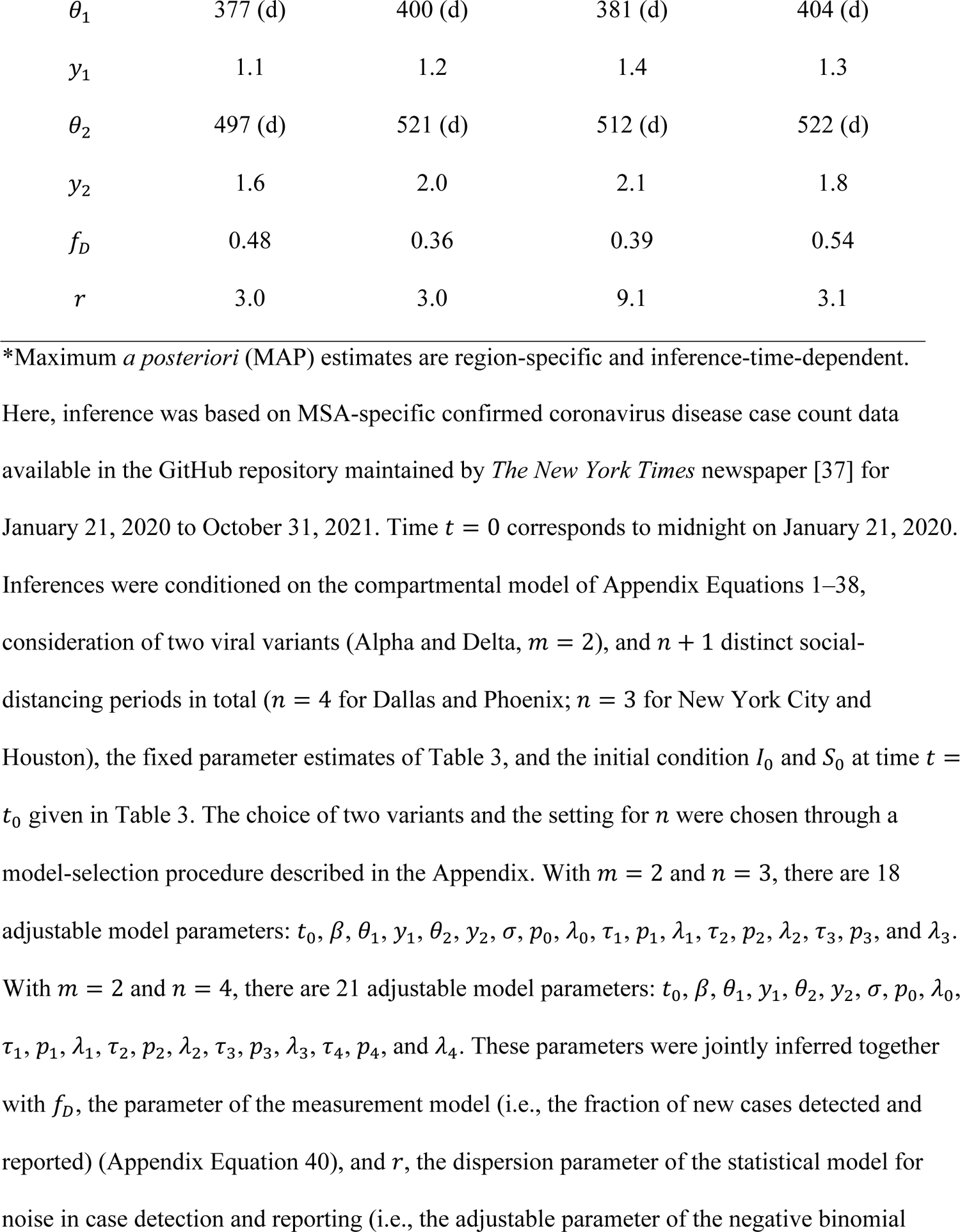
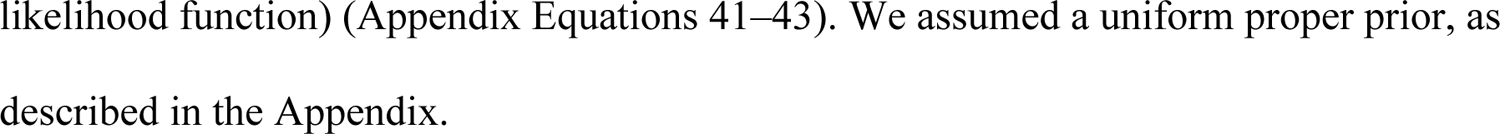
Model parameter values inferred for the Dallas, Houston, New York City, and Phoenix Metropolitan Statistical Areas (MSAs) on October 31, 2021 Here, inference was based on MSA-specific confirmed coronavirus disease case count data available in the GitHub repository maintained by *The New York Times* newspaper [37] for January 21, 2020 to October 31, 2021. Time *t* = 0 corresponds to midnight on January 21, 2020. Inferences were conditioned on the compartmental model of Appendix Equations 1–38, consideration of two viral variants (Alpha and Delta, *m* = 2), and *n* + 1 distinct social-distancing periods in total (*n* = 4 for Dallas and Phoenix; *n* = 3 for New York City and Houston), the fixed parameter estimates of Table 3, and the initial condition *I*_0_ and *S*_0_ at time *t* = *t*_0_ given in Table 3. The choice of two variants and the setting for *n* were chosen through a model-selection procedure described in the Appendix. With *m* = 2 and *n* = 3, there are 18 adjustable model parameters: *t*_0_, β, θ_1_, *y*_1_, θ), *y*_2_, σ, *p*_0_, λ_0_, τ_1_, *p*_1_, λ_1_, τ), *p*), λ), τ_*_, *p*_*_, and λ_*_. With *m* = 2 and *n* = 4, there are 21 adjustable model parameters: *t*_0_, β, θ_1_, *y*_1_, θ), *y*), σ, *p*_0_, λ_0_, τ_1_, *p*_1_, λ_1_, τ), *p*), λ), τ_*_, *p*_*_, λ_*_, τ_(_, *p*_(_, and λ_(_. These parameters were jointly inferred together with *f*_*D*_, the parameter of the measurement model (i.e., the fraction of new cases detected and reported) (Appendix Equation 40), and τ, the dispersion parameter of the statistical model for noise in case detection and reporting (i.e., the adjustable parameter of the negative binomial likelihood function) (Appendix Equations 41–43). We assumed a uniform proper prior, as described in the Appendix.

| Parameter | MAP estimate* | MAP estimate* | MAP estimate* for | MAP estimate* |
| --- | --- | --- | --- | --- |
|  | for Dallas | for Houston | New York City | for Phoenix |
|  | (Units) | (Units) | (Units) | (Units) |
| $t_0$ | 14.8 (d) | 11.2 (d) | 9.6 (d) | 14.0 (d) |
| $\beta$ | 0.30 (d<sup>-1</sup>) | 0.29 (d<sup>-1</sup>) | 0.37 (d<sup>-1</sup>) | 0.29 (d<sup>-1</sup>) |
| $\sigma$ | 65 (d) | 69 (d) | 68 (d) | 62 (d) |
| $p_0$ | 0.32 | 0.29 | 0.48 | 0.26 |
| $\lambda_0$ | 9.7 (d<sup>-1</sup>) | 7.5 (d<sup>-1</sup>) | 9.7 (d<sup>-1</sup>) | 6.6 (d<sup>-1</sup>) |
| $\tau_1$ | 139 (d) | 162 (d) | 143 (d) | 148 (d) |
| $p_1$ | 0.27 | 0.39 | 0.40 | 0.23 |
| $\lambda_1$ | 2.8 (d<sup>-1</sup>) | 0.1 (d<sup>-1</sup>) | 2.7 (d<sup>-1</sup>) | 2.5 (d<sup>-1</sup>) |
| $\tau_2$ | 165 (d) | 186 (d) | 177 (d) | 169 (d) |
| $p_2$ | 0.38 | 0.34 | 0.41 | 0.41 |
| $\lambda_2$ | 0.63 (d<sup>-1</sup>) | 0.53 (d<sup>-1</sup>) | 0.39 (d<sup>-1</sup>) | 0.47 (d<sup>-1</sup>) |
| $\tau_3$ | 259 (d) | 278 (d) | 262 (d) | 248 (d) |
| $p_3$ | 0.22 | 0.17 | 0.26 | 0.10 |
| $\lambda_3$ | 5.1 (d<sup>-1</sup>) | 3.2 (d<sup>-1</sup>) | 8.1 (d<sup>-1</sup>) | 5.2 (d<sup>-1</sup>) |
| $\tau_4$ | 264 (d) | - | - | 255 (d) |
| $p_4$ | 0.24 | - | - | 0.15 |
| $\lambda_4$ | 1.7 (d<sup>-1</sup>) | - | - | 3.2 (d<sup>-1</sup>) |
| $\theta_1$ | 377 (d) | 400 (d) | 381 (d) | 404 (d) |
| $y_1$ | 1.1 | 1.2 | 1.4 | 1.3 |
| $\theta_2$ | 497 (d) | 521 (d) | 512 (d) | 522 (d) |
| $y_2$ | 1.6 | 2.0 | 2.1 | 1.8 |
| $f_D$ | 0.48 | 0.36 | 0.39 | 0.54 |
| $r$ | 3.0 | 3.0 | 9.1 | 3.1 |

\*Maximum *a posteriori* (MAP) estimates are region-specific and inference-time-dependent.

**Table 2.** Descriptions of MSA-specific adjustable model parameters.

| Parameter | Description |
| --- | --- |
| $t_0$ | Start of local disease transmission |
| $\beta$ | Rate constant for disease transmission |
| $\sigma$ | Start of first social-distancing period |
| $p_0$ | Social-distancing setpoint for the period $t \in [\sigma, \tau_1)$ |
| $\lambda_0$ | Social-distancing eigenvalue paired with $p_0$ |
| $\tau_1$ | Start of second social-distancing period |
| $p_1$ | Social-distancing setpoint for the period $t \in [\tau_1, \tau_2)$ |
| $\lambda_1$ | Social-distancing eigenvalue paired with $p_1$ |
| $\tau_2$ | Start of third social-distancing period |
| $p_2$ | Social-distancing setpoint for the period $t \in [\tau_2, \tau_3)$ |
| $\lambda_2$ | Social-distancing eigenvalue paired with $p_2$ |
| $\tau_3$ | Start of fourth social-distancing period |
| $p_3$ | Social-distancing setpoint for the period $t \in [\tau_3, \tau_4)$ |
| $\lambda_3$ | Social-distancing eigenvalue paired with $p_3$ |
| $\tau_4$ | Start of fifth social-distancing period |
| $p_4$ | Social-distancing setpoint for the period $t \in [\tau_4, \infty)$ |
| $\lambda_4$ | Social-distancing eigenvalue paired with $p_4$ |
| $\theta_1$ | Alpha takeoff time |
| $y_1$ | Increased infectiousness of Alpha (relative to<br>ancestral strains) |
| $\theta_2$ | Delta takeoff time |
| $y_2$ | Increased infectiousness of Delta (relative to ancestral strains) |
| $f_D$ | Fraction of cases detected and reported |
| $r$ | Dispersion parameter of NB($r, q_i$)* |

**Table 3.**
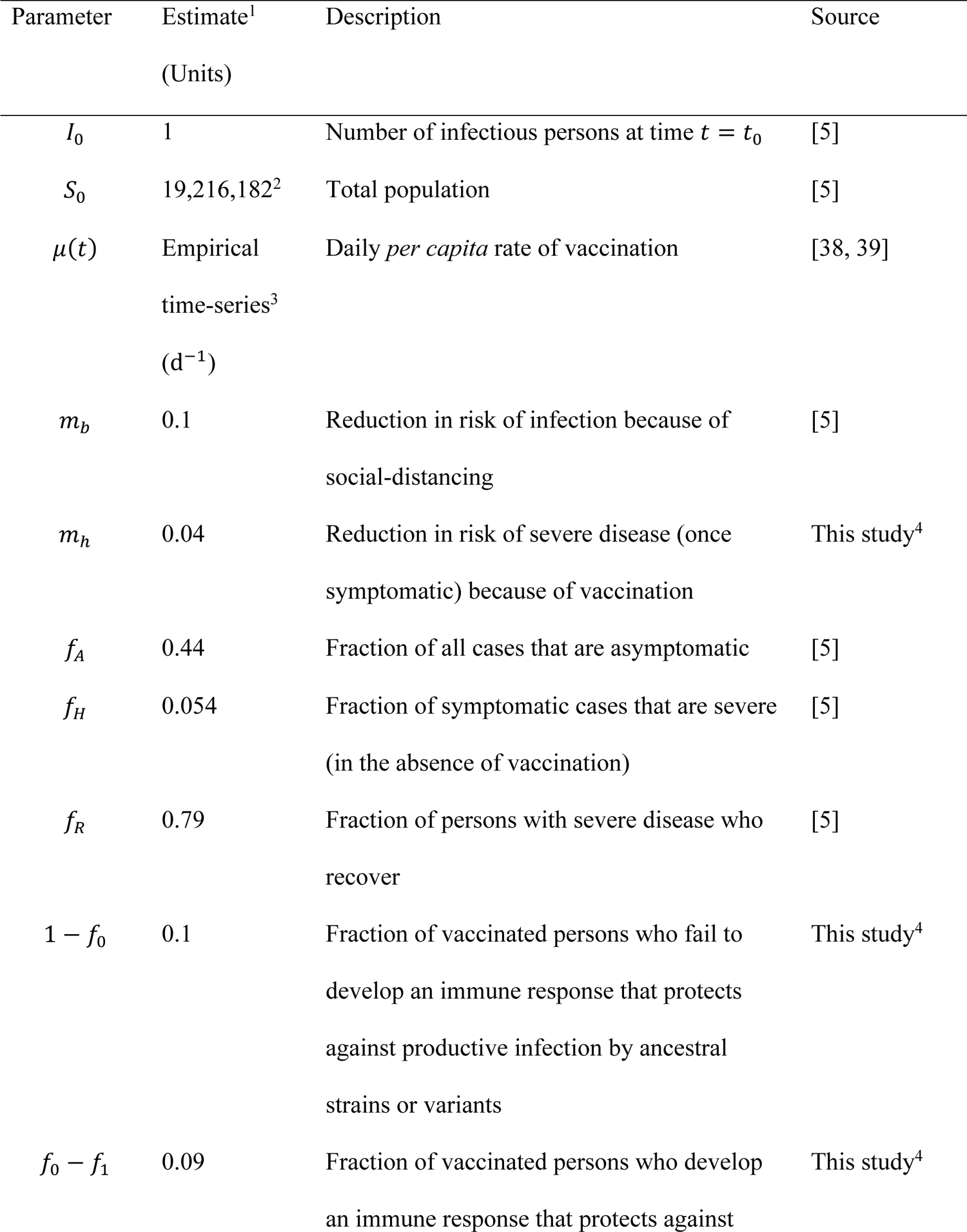
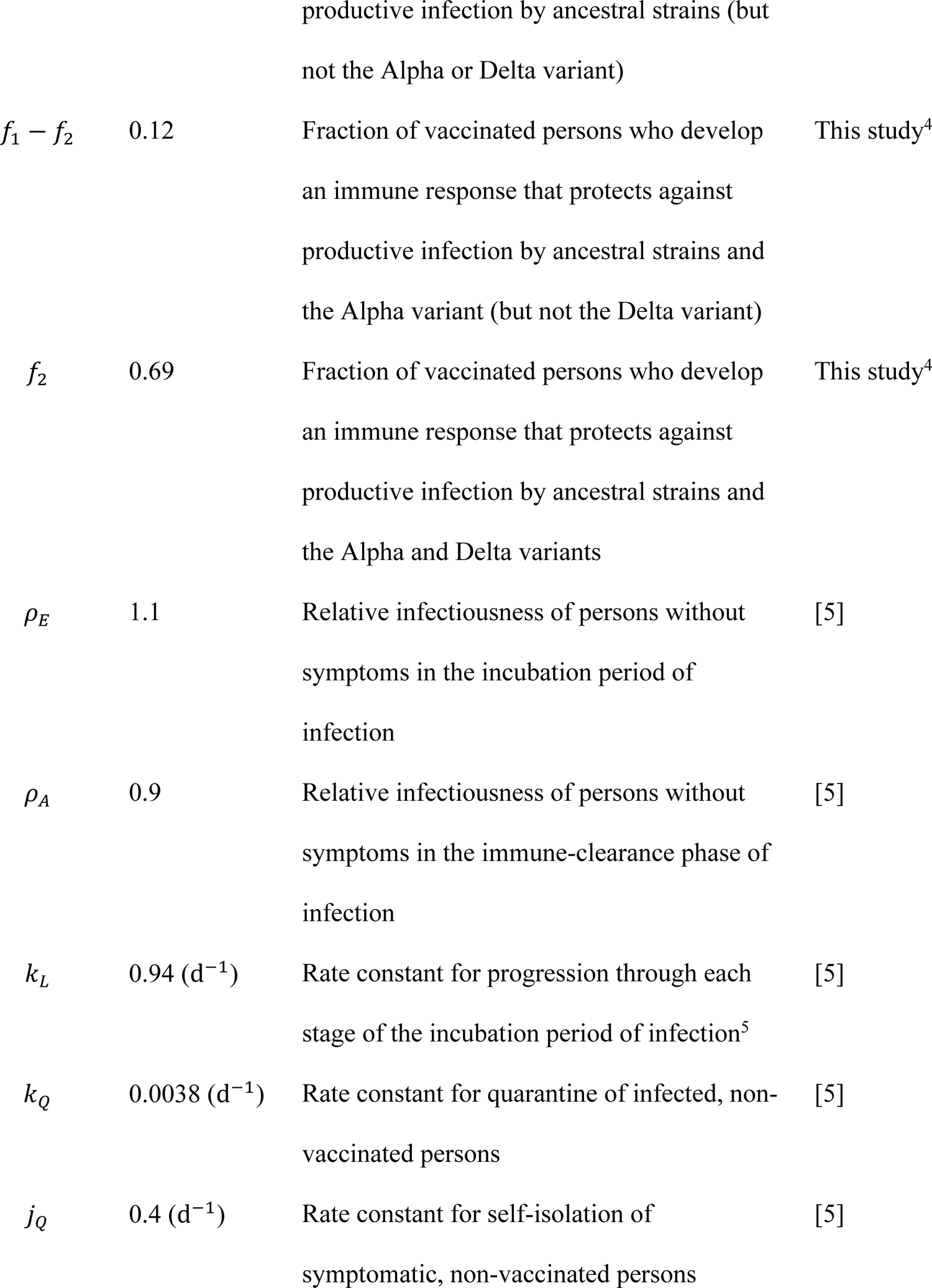
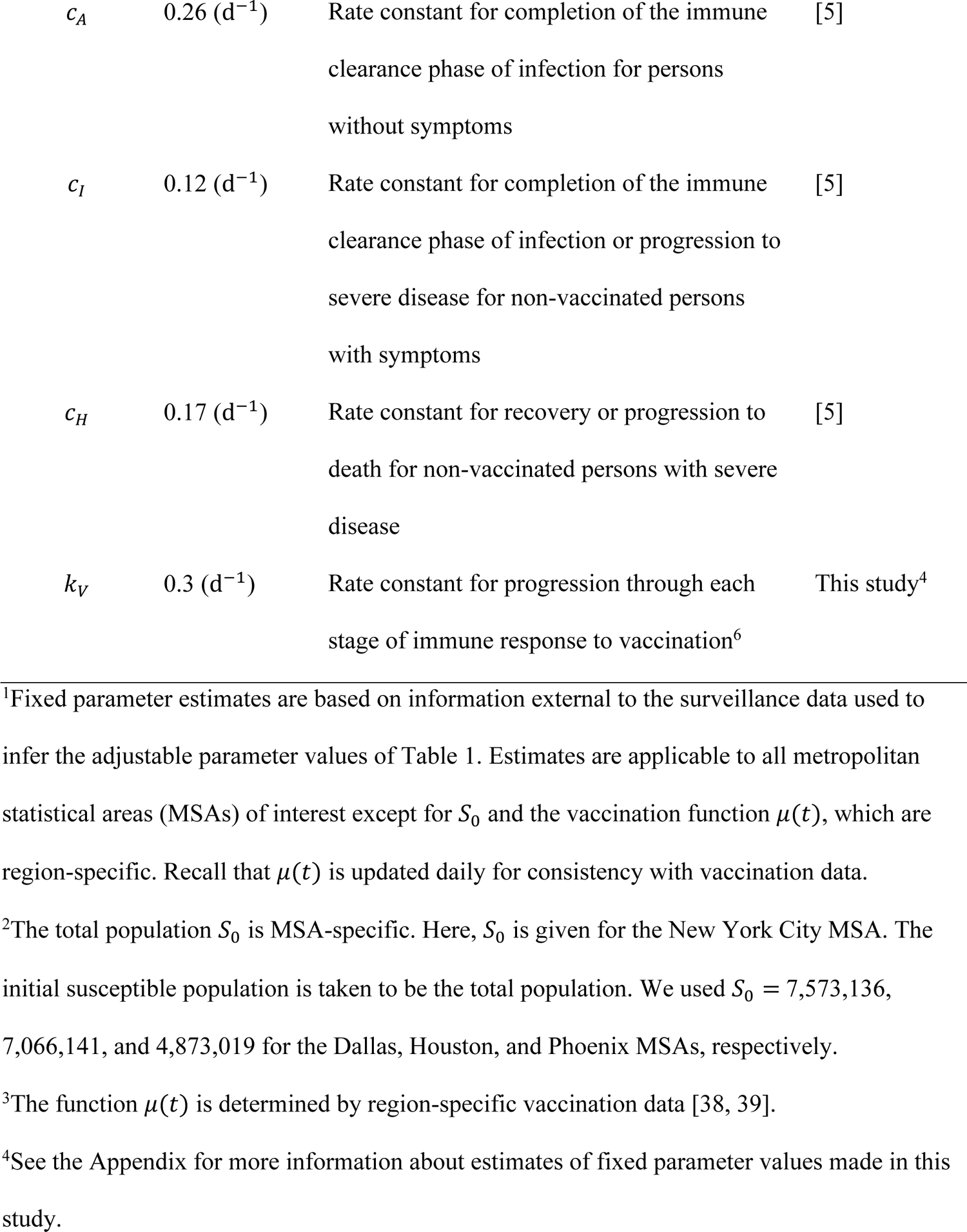
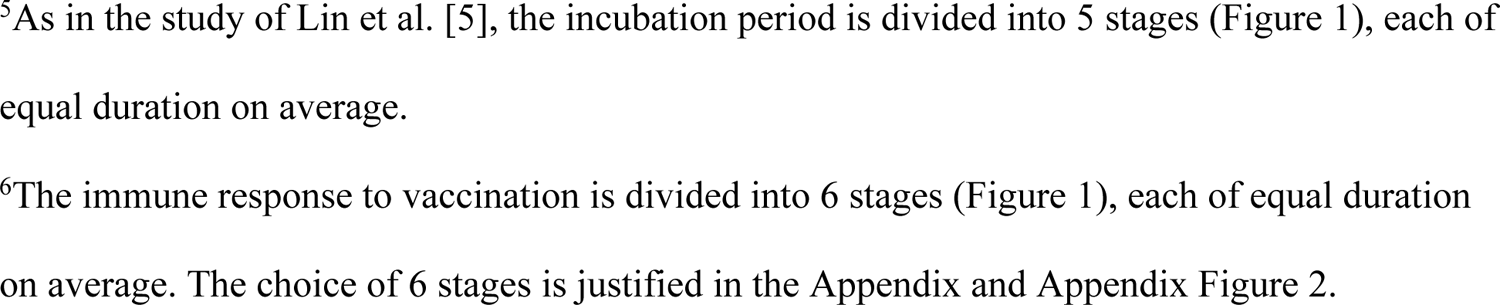
Fixed parameter estimates for each region-specific compartmental model.

| Parameter | Estimate<sup>1</sup><br>(Units) | Description | Source |
| --- | --- | --- | --- |
| $I_0$ | 1 | Number of infectious persons at time $t = t_0$ | [5] |
| $S_0$ | 19,216,182<sup>2</sup> | Total population | [5] |
| $\mu(t)$ | Empirical<br>time-series<sup>3</sup><br>(d<sup>-1</sup>) | Daily *per capita* rate of vaccination | [38, 39] |
| $m_b$ | 0.1 | Reduction in risk of infection because of social-distancing | [5] |
| $m_h$ | 0.04 | Reduction in risk of severe disease (once symptomatic) because of vaccination | This study<sup>4</sup> |
| $f_A$ | 0.44 | Fraction of all cases that are asymptomatic | [5] |
| $f_H$ | 0.054 | Fraction of symptomatic cases that are severe (in the absence of vaccination) | [5] |
| $f_R$ | 0.79 | Fraction of persons with severe disease who recover | [5] |
| $1 - f_0$ | 0.1 | Fraction of vaccinated persons who fail to develop an immune response that protects against productive infection by ancestral strains or variants | This study<sup>4</sup> |
| $f_0 - f_1$ | 0.09 | Fraction of vaccinated persons who develop an immune response that protects against productive infection by ancestral strains (but not the Alpha or Delta variant) | This study<sup>4</sup> |
| $f_1 - f_2$ | 0.12 | Fraction of vaccinated persons who develop an immune response that protects against productive infection by ancestral strains and the Alpha variant (but not the Delta variant) | This study<sup>4</sup> |
| $f_2$ | 0.69 | Fraction of vaccinated persons who develop an immune response that protects against productive infection by ancestral strains and the Alpha and Delta variants | This study<sup>4</sup> |
| $\rho_E$ | 1.1 | Relative infectiousness of persons without symptoms in the incubation period of infection | [5] |
| $\rho_A$ | 0.9 | Relative infectiousness of persons without symptoms in the immune-clearance phase of infection | [5] |
| $k_L$ | 0.94 (d<sup>-1</sup>) | Rate constant for progression through each stage of the incubation period of infection<sup>5</sup> | [5] |
| $k_Q$ | 0.0038 (d<sup>-1</sup>) | Rate constant for quarantine of infected, non-vaccinated persons | [5] |
| $j_Q$ | 0.4 (d<sup>-1</sup>) | Rate constant for self-isolation of symptomatic, non-vaccinated persons | [5] |
| $c_A$ | 0.26 (d<sup>-1</sup>) | Rate constant for completion of the immune clearance phase of infection for persons without symptoms | [5] |
| $c_I$ | 0.12 (d<sup>-1</sup>) | Rate constant for completion of the immune clearance phase of infection or progression to severe disease for non-vaccinated persons with symptoms | [5] |
| $c_H$ | 0.17 (d<sup>-1</sup>) | Rate constant for recovery or progression to death for non-vaccinated persons with severe disease | [5] |
| $k_V$ | 0.3 (d<sup>-1</sup>) | Rate constant for progression through each stage of immune response to vaccination<sup>6</sup> | This study<sup>4</sup> |

<sup>1</sup>Fixed parameter estimates are based on information external to the surveillance data used to infer the adjustable parameter values of Table 1. Estimates are applicable to all metropolitan statistical areas (MSAs) of interest except for $S_0$ and the vaccination function $\mu(t)$ , which are region-specific. Recall that $\mu(t)$ is updated daily for consistency with vaccination data.
<sup>2</sup>The total population $S_0$ is MSA-specific. Here, $S_0$ is given for the New York City MSA. The initial susceptible population is taken to be the total population. We used $S_0 = 7,573,136$ , 7,066,141, and 4,873,019 for the Dallas, Houston, and Phoenix MSAs, respectively.
<sup>3</sup>The function $\mu(t)$ is determined by region-specific vaccination data [38, 39].
<sup>4</sup>See the Appendix for more information about estimates of fixed parameter values made in this study.
<sup>5</sup>As in the study of Lin et al. [5], the incubation period is divided into 5 stages (Figure 1), each of equal duration on average.
<sup>6</sup>The immune response to vaccination is divided into 6 stages (Figure 1), each of equal duration on average. The choice of 6 stages is justified in the Appendix and Appendix Figure 2.

We extended the model of Lin et al. [5] by including 15 new populations and 28 new transitions. Briefly, new parts of the model can be described as follows. Vaccination is modeled by moving susceptible persons (in the *S*_*M*_ and *S*_*P*_populations) into the *V*_1_ population. Another consequence of vaccination is the movement of recovered unvaccinated persons (in the *R*_*U*_ population) into the *R*_*V*_ population. The rate of vaccination is changed as needed to match the empirical daily rate of vaccination. Recovered and susceptible persons have the same *per capita* probability of vaccination. Persons in *S*_*M*_ are mixing (i.e., not practicing social-distancing) and persons in *S*_*P*_ are practicing social-distancing (and thereby protected from infection to a degree). The series of transitions involving the populations *V*_1_, …, *V*_&_ was introduced to model the immune response to vaccination (i.e., the amount of time required for vaccination to induce neutralizing antibodies). With this approach, the time from vaccination to appearance of neutralizing antibodies is a random variable characterized by an Erlang distribution. Persons in *V*_1_, …, *V*_&_ may be infected. Persons in *V*_&_ transition to one of the following four populations: *S*_*V*,1_, …, *S*_*V*,(_. These populations represent persons with varying degrees of immune protection. Persons in *S*_*V*,1_ are not protected against productive infection (i.e., an infection that can be transmitted to others) by any viral strain. Persons in *S*_*V*,)_ are protected against productive infection by viral strains present before the emergence of Alpha but not Alpha or Delta. Persons in *S*_*V*,*_ are protected against productive infection by viral strains present before the emergence of Alpha and also Alpha but not Delta. Persons in *S*_*V*,(_ are protected against productive infection by all of the viral strains considered up to October 31, 2021. Vaccinated persons who become infected move into *E*_*V*_. The time spent in *E*_*V*_ corresponds to the length of the incubation period for vaccinated persons. The mean duration of the incubation period is taken to be the same for vaccinated and unvaccinated persons; however, as a simplification, for vaccinated persons, the time spent in the incubation period is taken to consist of a single stage and consequently is exponentially distributed (vs. Erlang distributed for unvaccinated persons). Non-quarantined exposed persons in populations *E*_*V*_ and *E*_+,*M*_ and *E*_+,*P*_ for *i* = 2, …, 5 are taken to be infectious. Persons exiting *E*_*V*_ leave the incubation period and enter the immune clearance phase of infection, during which they may be asymptomatic (*A*_*V*_) or symptomatic with mild disease (*I*_*V*_). Non-quarantined asymptomatic persons in populations *A*_*V*_, *A*_*M*_, and *A*_*P*_ are taken to be infectious. Persons in *A*_*V*_ are assumed to eventually recover (i.e., to enter *R*_*V*_). Persons with mild symptomatic disease may recover (i.e., enter *R*_*V*_) or experience severe disease, at which point they move to *H*_*V*_ (in hospital or isolated at home). Vaccinated persons have a diminished probability of severe disease in comparison to unvaccinated persons. Persons in *H*_*V*_ either recover (move to *R*_*V*_) or die from COVID-19 complications (move to *D*). For a person with severe disease, the probability of death is independent of vaccination status. We assume that vaccinated persons do not participate in social-distancing, quarantine, or self-isolation driven by symptom awareness.

The new compartments and transitions, which are highlighted in Figure 1, capture vaccination among susceptible persons, recovered persons and infected non-quarantined persons without symptoms at a time-varying *per capita* rate μ(*t*). The value of μ(*t*) changes daily for consistency with MSA-specific daily reports of completed vaccinations (Appendix Equation 37) from the COVID Act Now database and the *Democrat and Chronicle* COVID-19 vaccine tracker. The model also captures immune responses to vaccination yielding varying degrees of protection and consequences of breakthrough infection of vaccinated persons. Vaccine protection against transmissible infection was taken to be variant-dependent.

We introduced a dimensionless step function, denoted *Y*_θ_(*t*) (Appendix Equation 38), which multiplies the disease-transmission rate constant β to account for *m* variants. In this study, *m* = 2 (see Appendix Equations 1–4, 18–22, and 24). Thus, in the new model, the quantity *Y*_θ_(*t*)β (vs. β alone) characterizes disease transmissibility at time *t*. The step function *Y*_θ_(*t*) was initially assigned a value of *y*_0_ = 1, and the value of *Y*_1_(*t*) was allowed to increase at times *t* = θ_1_ and *t* = θ) (Appendix Equation 38). The disease transmission rate constant of the Alpha variant was considered by introducing a step increase from *y*_0_β ≡ 1β to *y*_1_β (with *y*_1_ > 1) at time *t* = θ_1_ (the Alpha takeoff time). Similarly, the disease transmission rate constant of the Delta variant was considered by introducing a step increase from *y*_1_β to *y*)β (with *y*) > *y*_1_) at time *t* = θ) > θ_1_ (the Delta takeoff time). We will refer to *y*_1_ and *y*) as the Alpha and Delta transmissibility factors, respectively.

As in the original model of Lin et al. [5], the extended model accounts for a series of *n* + 1 distinct social-distancing periods (an initial period and *n* additional periods). Social-distancing periods are characterized by two step functions: *P*_τ_(*t*) and Λ_τ_(*t*). The values of these functions change coordinately at a set of switch times τ = (σ, τ_1_, …, τ_*n*_) (Appendix Equations 35 and 36), where σ is the start time of the initial social-distancing period and τ_+_ is the start time of the *i*th social-distancing period after the initial social-distancing period. The values of *P*_τ_(*t*) and Λ_τ_(*t*) are zero before time *t* = σ. The value of *P*_τ_(*t*) defines the steady-state setpoint fraction of the susceptible population practicing social-distancing at time *t*, and the value of Λ_τ_(*t*) defines a time scale for establishment of the steady state. The value of Λ_τ_(*t*) is an eigenvalue equal to a sum of social-distancing rate constants [5]. The non-zero values of *P*_τ_(*t*) and Λ_τ_(*t*) are denoted *p*_0_, …, *p*_*n*_ and λ_1_, …, λ_*n*_. We assume that vaccinated persons do not practice social-distancing. Recall that we use the term “social-distancing” to refer to behaviors adopted to protect against infection. These behaviors are assumed to reduce the risk of infection by a factor *m*_*b*_.

### Parameters

As indicated in Tables 1 and 2, we used MSA-specific case reporting data available up to October 31, 2021 to infer MSA-specific values for parameters characterizing the start time of the local epidemic (*t*_0_), local disease transmissibility of ancestral viral strains (β), local social-distancing dynamics (σ, λ_1_, *p*_1_, τ_+_, λ_+_ and *p*_+_, for *i* = 1, …, *n*), local emergence of variants (θ_1_, *y*_1_, θ), *y*)), the local rate of new case detection (*f*_*D*_), and noise in local case detection and reporting (τ). Values for other parameters were fixed (Table 3); inferences are conditioned on these fixed parameter estimates. There are 18 fixed parameters taken to be applicable for all MSAs. The total regional population *S*_2_, which is taken to be fixed, was set on the basis of census data. The real-time *per capita* vaccination rate μ(*t*), a piecewise linear function, was set for consistency with the current empirical *per capita* rate of vaccination [38]. We adopted the fixed parameter estimates of Lin et al. [5]. New fixed parameter estimates made in this study for *m*_2_, *f*_0_, *f*_1_, *f*_2_, and *k*_*V*_ are explained in the Appendix. The *m*_2_ parameter characterizes vaccine protection against severe disease, the *f*_0_, *f*_1_, and *f*) parameters account for differential vaccine effectiveness against the three viral strains considered in this study (ancestral, Alpha, and Delta), and the *k*_*V*_ parameter characterizes the waiting time between vaccination and the acquisition of vaccine-induced immunity. Our model does not account for gradual loss of immunity over time. We let μG(*t*) = μ_+_ for times *t* throughout the *i*th day after January 21, 2020 (Appendix Equation 37), where μ_+_ is the fraction of the local population reported to complete vaccination over the 1-d surveillance period [38]. We then defined μ(*t*) as the piecewise linear interpolant to μG(*t*). In summary, for a given inference, the number of adjustable parameters was 2*m* + 3*n* + 4, where *m* is the number of variants under consideration (*m* = 2 in this study) and *n* is the number of distinct social-distancing periods being considered beyond an initial social-distancing period. The setting for *n* was determined through a model-selection procedure described in the Appendix.

### Auxiliary Measurement Model

We assumed that state variables of the compartmental model (Figure 1, Appendix Table 1) are related to the expected number of new cases reported on a given calendar date through an auxiliary measurement model (Appendix Equations 39 and 40). The measurement model has one parameter: *f*_*D*_, the region-specific fraction of new symptomatic infections detected. Thus, *f*_*D*_ ∈ [0, 1]. As a simplification, we considered *f*_*D*_ to be time-invariant. This simplification means that we assumed, for example, that case detection was neither limited nor strongly influenced by testing capacity, which varied over time. This assumption is reasonable if, for example, case detection is mainly determined by presentation for testing and, moreover, the motivations and societal factors that influence presentation remained roughly constant over the period of interest. One can also interpret *f*_*D*_ as the time-averaged case detection rate. The measurement-model parameter *f*_*D*_was inferred jointly with the adjustable model parameters and the likelihood parameter τ (see below).

### Statistical Model for Noise in Case Detection and Reporting

We assumed that noise in new case detection and reporting on the *i*th day after January 21, 2020 is captured by a negative binomial distribution NB(τ, *q*_i_) centered on *I*(*t*_+_, *t*_+31_), the expected number of new cases detected over the *i*th day after January 21, 2020 as given by the compartmental model and the auxiliary measurement model (Appendix Equations 1–40). These and other assumptions led to the likelihood function used in inference (Appendix Equations 41– 43). We determined the probability parameter *q*_+_ in NB(τ, *q*_+_) using Appendix Equation 43; the dispersion parameter τ was taken to be a time-invariant adjustable parameter applicable for all days of case reporting. The likelihood parameter τ was inferred jointly with the adjustable model parameters and the adjustable measurement-model parameter *f*_*D*_.

### Computational Procedures

We determined the intervals of the step functions *Y*_θ_(*t*), *P*_τ_(*t*), and Λ_τ_(*t*) (i.e., θ and τ) using a model-selection procedure described in the Appendix. Simulations and Bayesian inferences were performed as previously described [5–7] and in the Appendix. Files needed to reproduce inferences using the software package PyBioNetFit [40] are available online [41]. The files include case data, vaccination data, and diagnostic plots related to Bayesian inference using Markov chain Monte Carlo (MCMC) sampling, including trace plots [41]. Summary diagnostics characterizing the sampling for each MSA are given in Table 4. Briefly, we computed the stable Gelman-Rubin statistic using the methodology of Vats and Knudson [42]. An advantage of this statistic (over the original Gelman-Rubin statistic) is that only one Markov chain is required in sampling. In addition, the stable Gelman-Rubin statistic and effective sample size (ESS) have a one-to-one relationship.

**Table 4.** Multivariate potential scale reduction factors (PSRFs) and multivariate effective sample sizes (ESSs) corresponding to the stable Gelman-Rubin statistic for Markov chains generated using PyBioNetFit for the Dallas, Houston, New York City, and Phoenix Metropolitan Statistical Areas (MSAs)

| MSA | Multivariate<br>PSRF<sup>1</sup> | Multivariate<br>ESS<sup>1</sup> |
| --- | --- | --- |
| Dallas | 1.0024 | 211.6 |
| Houston | 1.0026 | 193.2 |
| New York City | 1.0023 | 212.5 |
| Phoenix | 1.0033 | 152.9 |

<sup>1</sup>Computed using the methodology of Vats and Knudson [42].

## RESULTS

### Explanatory power of the model

As illustrated in Figures 2A, 3A, 4A, and 5A for the Dallas, Houston, New York City, and Phoenix MSAs respectively, the new model is able to explain surveillance case data over the period starting on January 21, 2020 and ending on October 31, 2021. The surveillance case data—daily reports of newly detected COVID-19 cases—for each of the 4 MSAs of interest largely lie within the 95% credible interval of the posterior predictive distribution for new case detection, which indicates that each regional model has explanatory power for the period of interest. Parameter estimates are summarized in Tables 1–3. Table 1 provides region-specific maximum *a posteriori* (MAP) estimates obtained through Bayesian inference enabled by Markov chain Monte Carlo (MCMC) sampling. Table 4 provides metrics that measure the quality of MCMC sampling. The diagnostic results of Table 4 indicate that sampling was acceptable, because all multivariate potential scale reduction factors (PSRFs) are very close to 1 and all multivariate effective sample sizes (ESSs) are at least 100. Diagnostic plots are provided online [41].

**Figure 2.**
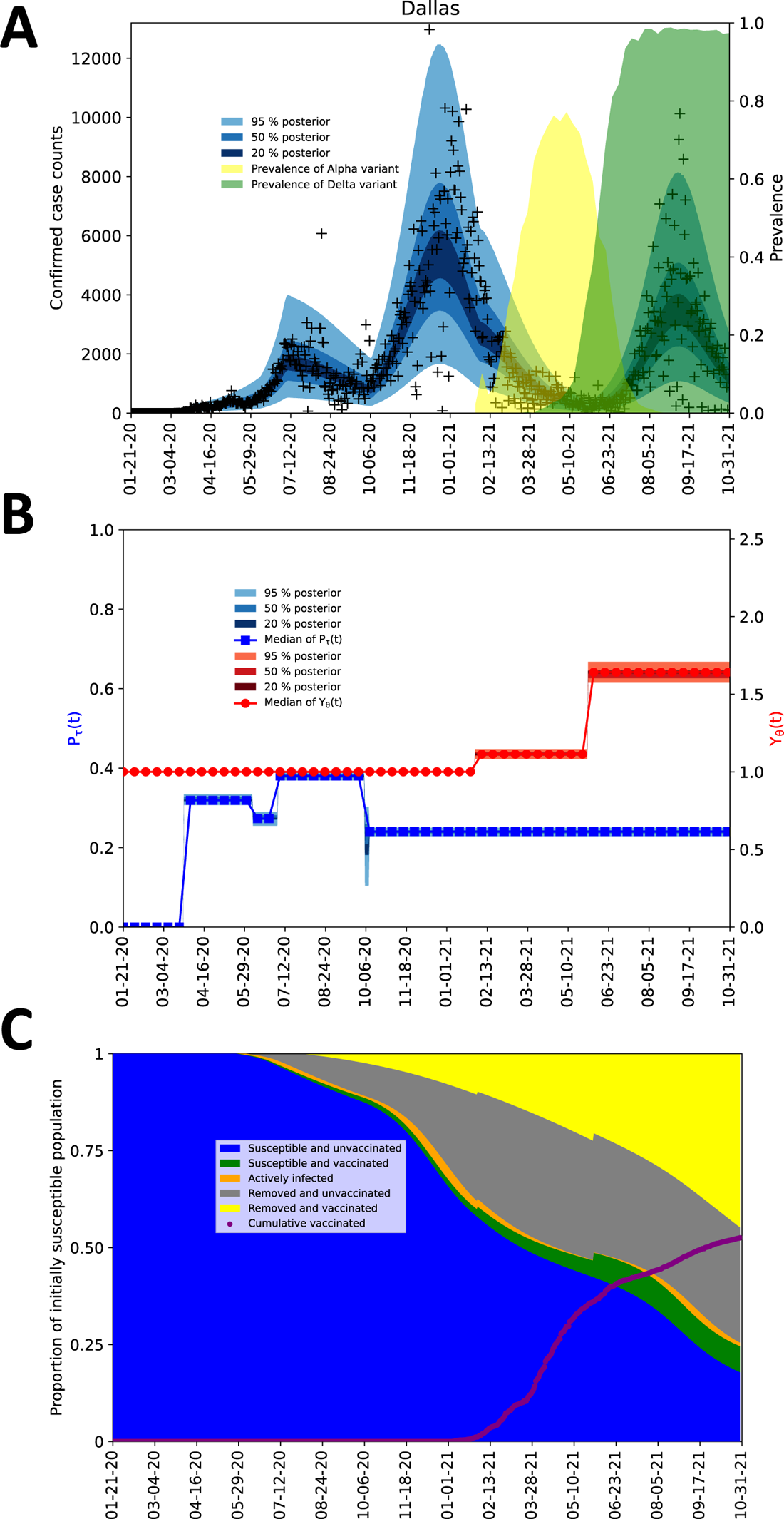
Inference results obtained for the MSA surrounding Dallas using regional surveillance data—daily reports of new COVID-19 cases—available for January 21, 2020 to October 31, 2021. A) Credible intervals of the time-dependent posterior predictive distribution for detected and reported new cases are shown. The monochrome bands from innermost to outermost indicate the 20%, 50%, and 95% credible intervals. This colored region all together indicates the 95% credible interval and can be expected to cover approximately 95% of the data. Empirical case reports are indicated by black symbols. Alpha prevalence is indicated with a shaded light-yellow background. Delta prevalence is indicated with a shaded light-green background. B) The social-distancing stationary setpoint is given by *P*_τ_(*t*) and the variant transmissibility factor is given by *Y*_θ_(*t*). Note that the values of *P*_τ_(*t*) and *Y*_θ_(*t*) are dimensionless. Credible intervals corresponding to 1000 samples from the time-dependent posterior predictive distributions are shown for *P*_τ_(*t*) and *Y*_θ_(*t*). The curve corresponding to *Y*_θ_(*t*) is monotonically increasing with an initial value of 1. The curve corresponding to *P*_τ_(*t*) is decreasing from left to right after the start of social-distancing. C) Inferred changes in the distribution of persons amongst five selected subpopulations over the course of the local COVID-19 epidemic. The five populations sum to a constant, *S*_0_, the total population. Results shown here are based on the parameter values given in Tables 1 and 3. The five populations are defined as follows: the population of susceptible unvaccinated persons (blue area) is given by *S*_*M*_ + *S*_*P*_, the population of susceptible vaccinated persons (green area) is given by ∑^&^ *V*_+_ + *S*_*V*,1_ + *U*_θ_ (*t*)*S*_*V*,)_ + *U*_θ_ (*t*)*S*_*V*,*_, the population of actively infected persons (orange area) is given by *H_U_ + H_V_ + E_V_ + Σ^5^_i=1_(E_i,m_ + E_i,p_ + E_i,Q_ + Σ_x∈{M,P,Q,V|_(A_x_ + I_x_)*, the population of removed unvaccinated persons (gray area) is given by *R*_*U*_ + *D*, and the population of removed vaccinated persons (yellow area) is given by *R*_*V*_ + (1 − *U*_1_(*t*))*S*_*V*,)_ + (1 − *U*_2_(*t*))*S*_*V*,*_ + *S*_*V*,(_. Except for *U*_1_(*t*) and *U*_2_(*t*), the terms in the above definitions refer to state variables of the compartmental model of Figure 1 and Appendix Figure 1, which are defined in Appendix Table 1. *U*_1_(*t*) and *U*_2_(*t*) are unit step functions (Appendix Equation 33 and 34), which change value from 0 to 1 at time *t* = θ_1_ and *t* = θ), respectively. Recall that θ_1_ and θ) are the Alpha and Delta takeoff times. The sum ∑_+_(μ_+_ × 1 d) (purple dots), which is the empirical cumulative number of completed vaccinations, is shown as a function of time *t*.

### Quantification of the impacts of non-pharmaceutical interventions and emergence of variants

Each regional model (parameterized to reproduce MSA-specific case reports) provides insight into the impacts of social-distancing behaviors and the emergence of the Alpha and Delta variants; compare panel A and the corresponding panel B in Figures 2–5. For example, as can be seen in Figure 4A, the New York City MSA experienced four notable surges in disease incidence over the period of interest. Figure 4B suggests that the first surge ended because of adoption of social-distancing behaviors, the second surge occurred because of relaxation of social-distancing behaviors, the third surge was caused by Alpha, and the fourth surge was caused by Delta.

**Figure 3.**
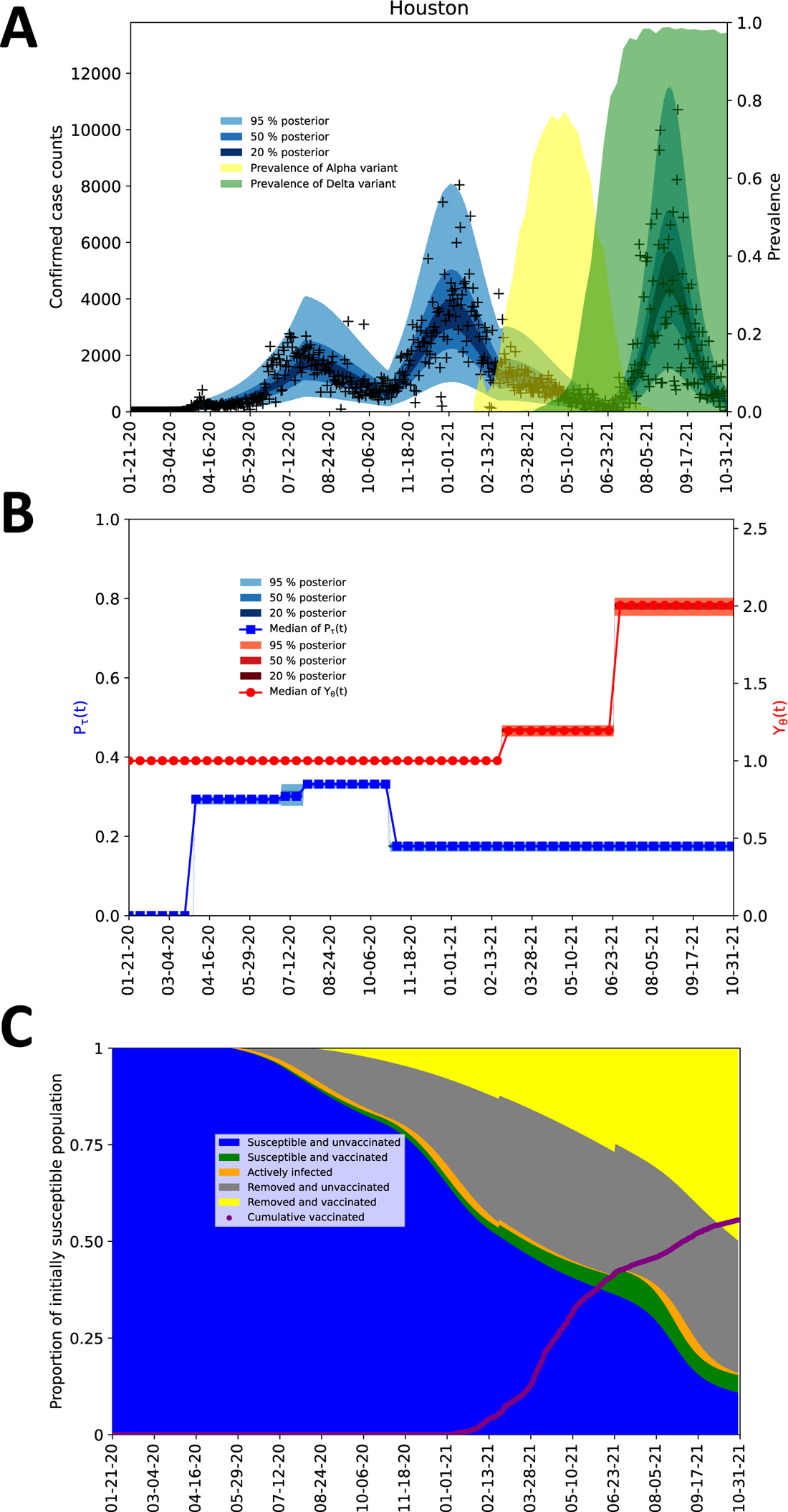
Inference results obtained for the MSA surrounding Houston using regional surveillance data—daily reports of new COVID-19 cases—available for January 21, 2020 to October 31, 2021. It should be noted that four anomalous (negative) empirical case counts are not shown in the plot. A case count of 14,300 cases on September 21, 2020 is not shown in the plot. See the caption of Figure 2 for additional information.

**Figure 4.**
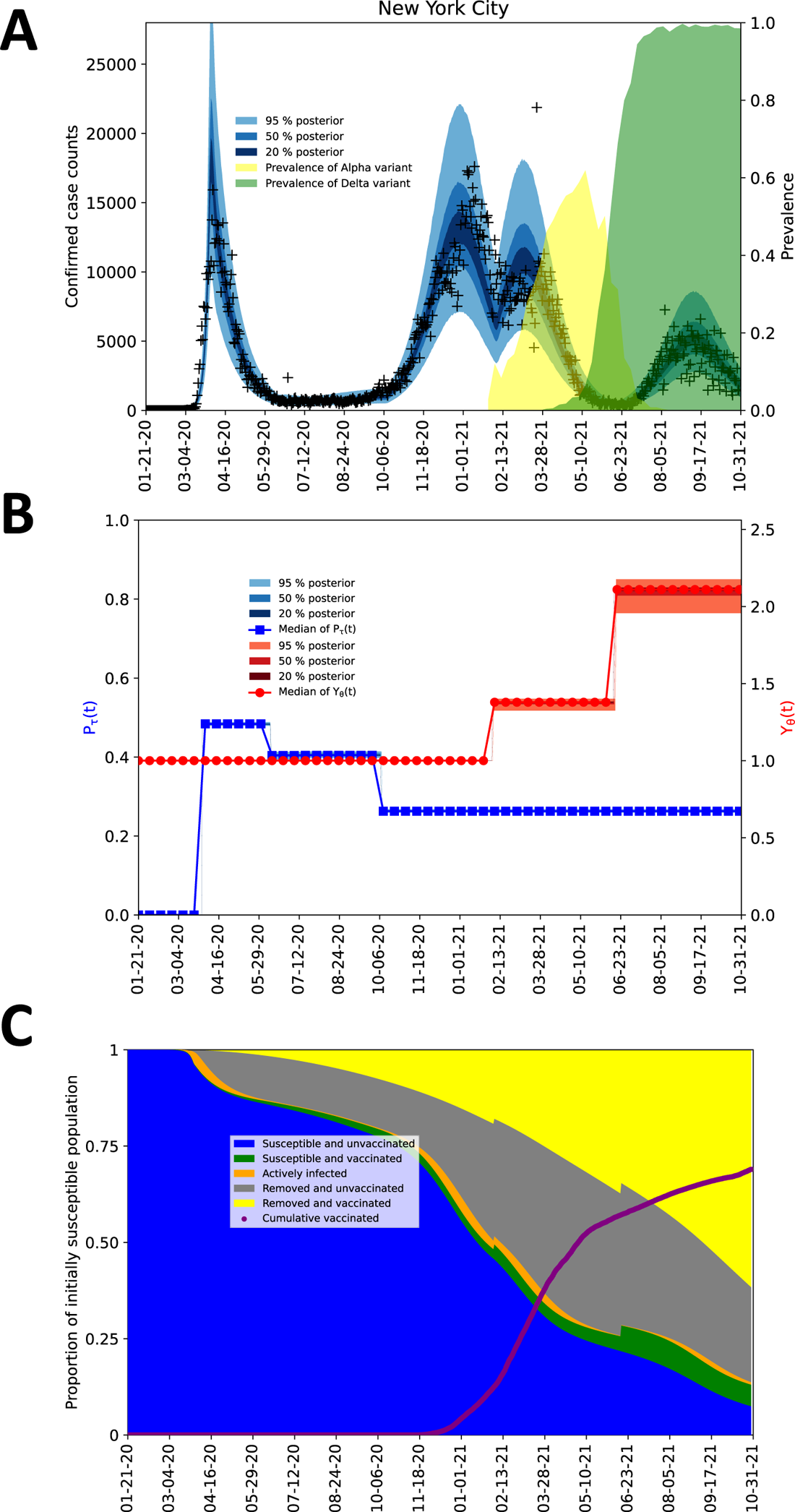
Inference results obtained for the MSA surrounding New York City using regional surveillance data—daily reports of new COVID-19 cases—available for January 21, 2020 to October 31, 2021. It should be noted that a single anomalous (negative) empirical case count is not shown in the plot. See the caption of Figure 2 for additional information.

**Figure 5.**
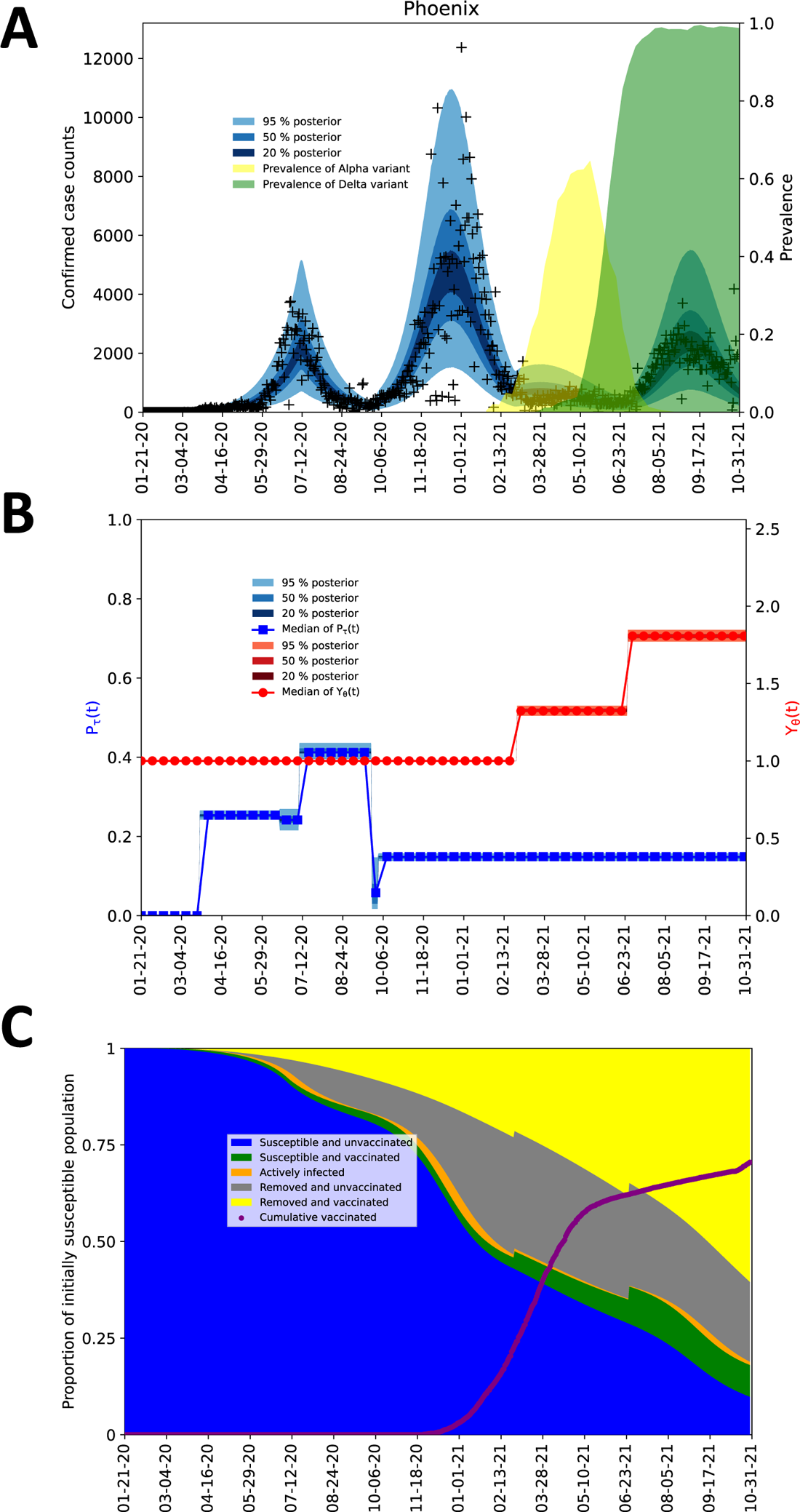
Inference results obtained for the MSA surrounding Phoenix using regional surveillance data—daily reports of new COVID-19 cases—available for January 21, 2020 to October 31, 2021. It should be noted that two anomalous (negative) empirical case counts are not shown in the plot. See the caption of Figure 2 for additional information.

Interestingly, in other MSAs, Alpha had relatively little impact on disease incidence (compare Figures 4A and 4B to Figures 2A and 2B, 3A and 3B, and 5A and 5B). These differences are attributable to various factors, including the lower contagiousness of COVID-19 in Dallas, Houston, and Phoenix relative to New York City (see entries for β in Table 1), the later arrival of Alpha in Houston and Phoenix relative to New York City, and further progress of mass vaccination in Phoenix relative to New York City. Alpha takeoff, as well as Delta takeoff, is tracked locally in panel B (rightmost vertical axis) of Figures 2–5, and vaccination progress is tracked locally in panel C of Figures 2–5.

### Impacts of vaccination

On the basis of region-specific parameterizations, we estimated the immune and susceptible fractions of each MSA population, as well as the fractions that achieved immunity through infection and/or vaccination (see panel C in Figures 2–5). Each of these panels shows the time evolution of five different populations in an MSA. In each MSA, only a minority of the population remained susceptible to infection (with Delta) on October 31, 2021, with a sizable fraction of the susceptible population being protected to a degree against severe disease by having completed vaccination.

### Estimates of transmissibility factors and takeoff times of Alpha and Delta

Our inferences provide quantitative insights into the transmissibility factors of Alpha and Delta (*y*_1_ and *y*)) and their takeoff times (θ_1_ and θ)) in each of the 4 MSAs of interest. Figure 6 shows the marginal posteriors of *y*_1_, *y*), θ_1_, and θ), which were found on the basis of surveillance data for each MSA (daily case counts) reported between January 21, 2020 and October 31, 2021. Maximum *a posteriori* (MAP) estimates and 95% credible intervals for θ_1_ and θ) for each MSA are also shown in Figure 6. The MAP estimates for *y*_1_ range from 1.1 (for the Dallas MSA) to 1.4 (for the New York City MSA). The MAP estimates for *y*) range from 1.6 (for the Dallas MSA) to 2.1 (for the New York City MSA). The MAP estimates for θ_1_ range from February 1, 2021 (for the Dallas MSA) to February 28, 2021 (for the Phoenix MSA). The MAP estimates for θ) range from June 2, 2021 (for the Dallas MSA) to June 26, 2021 (for the Houston and Phoenix MSAs). The estimated takeoff times are consistent with regional genomic surveillance data [43], which are summarized by the shaded regions of panel A in Figures 2–5.

**Figure 6.**
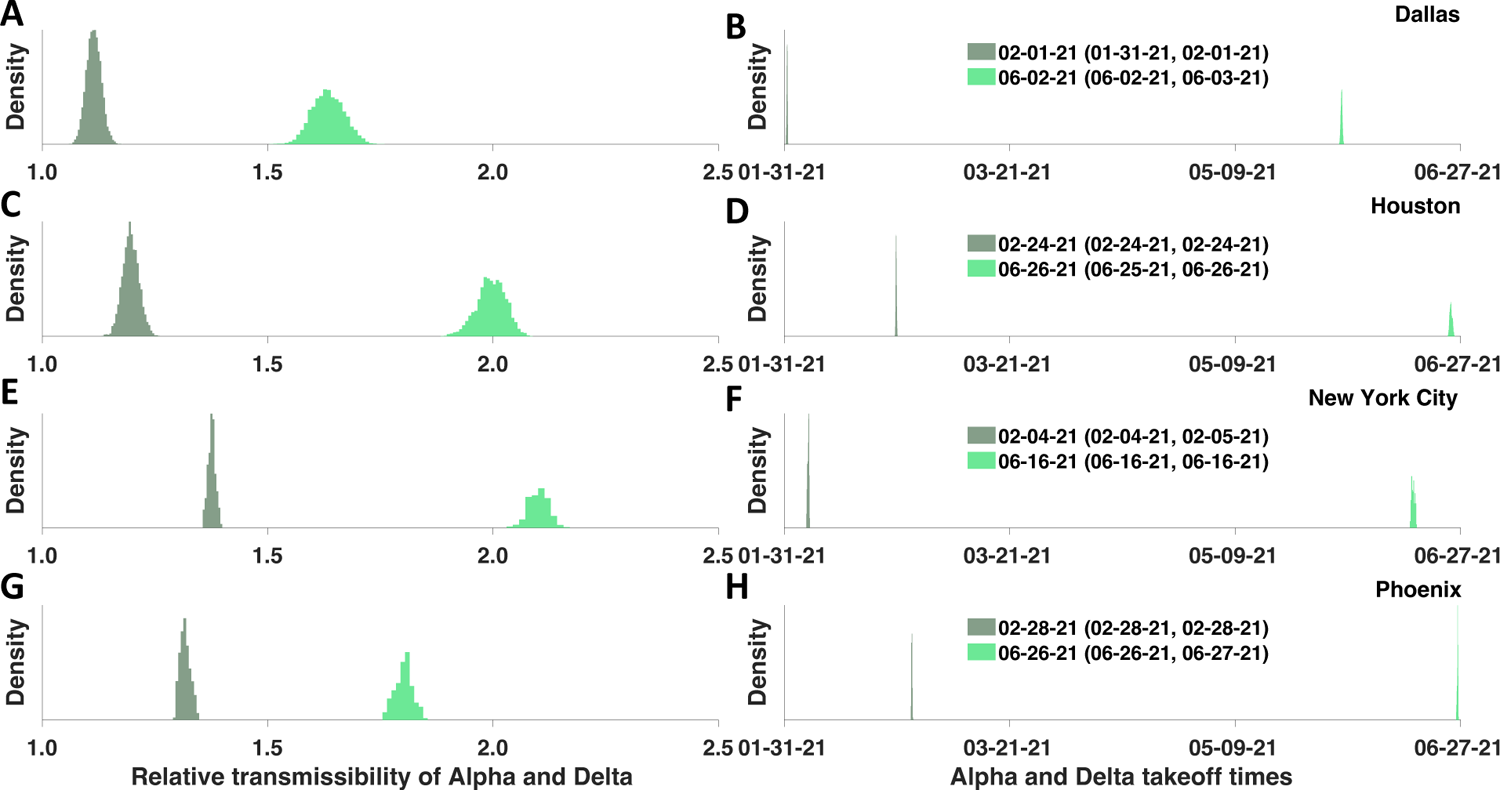
Marginal posterior distributions of transmissibility factors for Alpha in dark-green and Delta in light-green (left panels) and takeoff times for Alpha and Delta (right panels) in four MSAs centered on (**A,B**) Dallas, (**C,D**) Houston, (**E,F**) New York City, and (**G,H**) Phoenix. Inferences are based on daily reports of new cases from January 21, 2020 to October 31, 2021. For each of the right panels, the 95% credible intervals for Alpha and Delta takeoff times are indicated in parentheses, and the MAP estimate for a given region is indicated to the left of the credible interval.

## DISCUSSION

We extended a model for COVID-19 transmission dynamics that already incorporated time-varying changes in non-pharmaceutical interventions [5] to include the effects of vaccination and new variants. This model together with its region-specific parameterizations based on case data available through October 31, 2021 provide quantitative insights into the relative infectiousness of SARS-CoV-2 variants Alpha (lineage B.1.1.7) and Delta (lineage B.1.617.2). The increased transmissibility of Alpha and Delta in comparison to ancestral strains is characterized by the marginal posteriors for the transmissibility factors *y*_1_ and *y*) shown in Figure 6 (panels A, C, E, and G). The maximum *a posteriori* (MAP) estimates of the transmissibility factors were similar across the four metropolitan statistical areas (MSAs) of interest (centered on Dallas, Houston, New York City, and Phoenix). The averages of our *y*_1_ and *y*) MAP estimates indicate that Alpha was 1.2x more infectious than ancestral strains, whereas Delta was 1.9x more infectious (corresponding to Delta being approximately 50% more infectious than Alpha). These estimates are consistent with estimates provided in other studies [18–23]. Using a formula for the basic reproduction number *R*_0_ derived earlier [7] and parameter estimates of Tables 1 and 3, we obtain an *R*_1_ estimate of 12 for Delta in the New York City MSA, which places this SARS-CoV-2 variant among the more infectious viruses known.

We also obtained estimates of precisely when sustained transmission of Alpha and Delta began in each of the four geographically distinct MSAs. In earlier work, takeoff times for Alpha and Delta were estimated only for regions outside the US (e.g., Denmark [44] and Mexico [45]), for regions in the US different from those considered here (e.g., New England [46] and two MSAs in Northern California [47]), and for the entire US [48]. The takeoff times for the four MSAs in the present study are characterized by the marginal posteriors for θ_1_ and θ) shown in Figure 6 (panels B, D, F, and H). The estimated takeoff times are consistent with the observed prevalence of Alpha and Delta sequences detected in regional genomic surveillance [43], as can be seen by comparing the two shaded regions in panel A of Figures 2–5 against the changes in transmissibility *Y*(*t*) depicted in the corresponding panel B of Figures 2–5. It should be noted that the case data shown in Figures 2–5 are from the MSAs of interest (through aggregation of county-level data), whereas the genomic surveillance data are from larger regions [43].

Our study has notable limitations, starting with the obvious uncertainties related to model assumptions and fixed parameter estimates, which are discussed in some detail in the Appendix. For example, our model assumes a constant rate of case detection and neglects gradual loss of sterilizing immunity over time. This latter simplifying assumption is supported by a study indicating that protection against re-infection from pre-Omicron variants is significant and remains high even after 40 weeks [49]. Moreover, we caution that the inference jobs performed in this study were challenging because of the relatively high-dimensional parameter spaces involved (in comparison to typical inferences involving an ODE model-constrained likelihood function). Diagnostics indicate good sampling (Table 4, [41]), but we cannot be entirely certain that the samples obtained fully characterize the parameter posteriors of interest. It can be seen in trace plots [41] that mixing across parameters was not uniform, and moreover, in some cases, there are indications of trends (manifesting as a trace plot that lacks the appearance of a horizontally extended, fuzzy caterpillar). The likely cause of these trace plots is poor local performance of the proposal kernel, which was optimized in the adaptive sampling scheme for global performance. The consequences of poor mixing may include biased parameter and uncertainty estimates. Another concern is that the model incorporates redundant disease-incidence surge mechanisms. In the model, an increase in viral infectiousness caused by the emergence of a new variant can be mimicked, to some extent, by a relaxation of social-distancing, and vice versa. For the MSAs considered here, inferred social-distancing levels were low at the time of Alpha and Delta emergence, so the inferred transmissibility factors probably reflect, at least mostly, changes in intrinsic viral infectiousness.

It is known that viral transmissibility does not depend only on viral features but also on population features [7]. This study looked at multiple regions to ascertain whether population features varied significantly. Our analysis indicates that population features influencing transmission did not vary dramatically across these similar urban regions because we obtained similar region-specific estimates for the Alpha and Delta transmissibility factors. The Alpha transmissibility factors for the Dallas, Houston, New York City, and Phoenix MSAs were 1.1x, 1.2x, 1.4x, and 1.3x, respectively. the Delta transmissibility factors for these MSAs were 1.6x, 2.0x, 2.1x, and 1.8x, respectively.

We show that it is possible to gain insights into variant dynamics through a mechanistic modeling approach. This study provides an example for modelers interesting in understanding the impacts of a mass vaccination campaign and emergence of variants with altered transmissibility during an epidemic of an aerosol-transmitted respiratory disease similar to COVID-19.

In summary, this report provides estimates of Alpha and Delta transmissibility for specific regions within the US as well as their takeoff times.

## Data Availability

Files needed to reproduce the Bayesian inference results and MCMC sampling diagnostic plots are available online at https://github.com/lanl/PyBNF/tree/master/examples/Vax_and_Variants

## ACKNOWLEDGMENTS

W.S.H., Y.T.L., and A.M. were supported by the LDRD program at Los Alamos National Laboratory (project 20220268ER). Y.C., Z.H., E.F.M., J.N., K.E.N., and R.G.P. were supported by a grant from the National Institute of General Medical Sciences of the National Institutes of Health (grant R01GM111510). A.M. was supported by the 2020 Mathematical Sciences Graduate Internship program, which is sponsored by the Division of Mathematical Sciences of the National Science Foundation, and the Center for Nonlinear Studies at Los Alamos National Laboratory. Computational resources used in this study included Northern Arizona University’s Monsoon computer cluster, which is funded by Arizona’s Technology and Research Initiative Fund, and the FARM computer cluster at the University of California, Davis. Y.C. thanks Song Chen (University of Wisconsin, La Crosse, Wisconsin, USA) for technical assistance.

## APPENDIX

### Imputation of Missing Daily Case Counts

By October 31, 2021, many regions in the US were not reporting new detected COVID-19 cases on a strictly daily basis. When one or more daily case counts were not available, we imputed daily case counts on the basis of a linear fit to the two nearest available cumulative case counts. This approach has the effect of evenly distributing case counts across the days for which daily reports are unavailable.

### Equations of the Compartmental Model

The compartmental model, which is illustrated in Figure 1 and Appendix Figure 1, consists of the following 40 ordinary differential equations (ODEs):

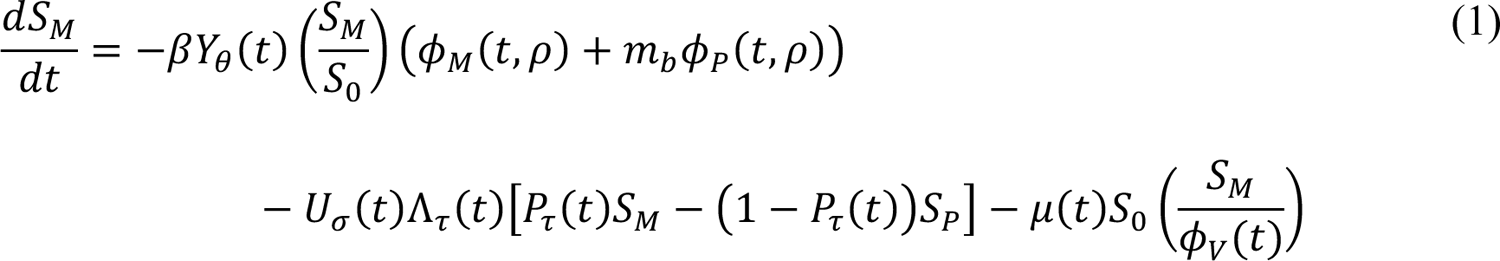

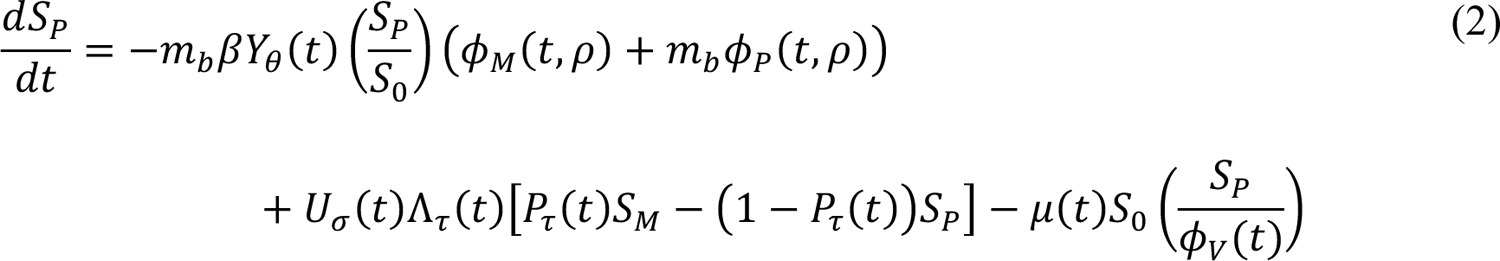

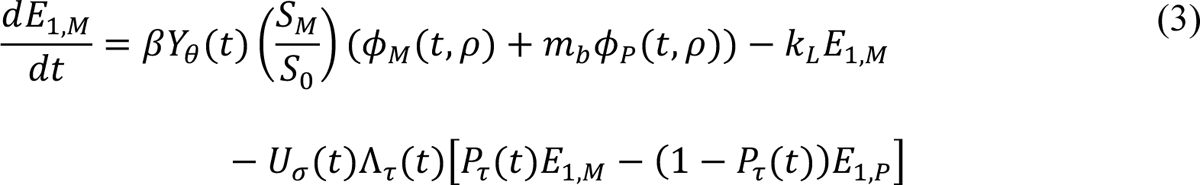

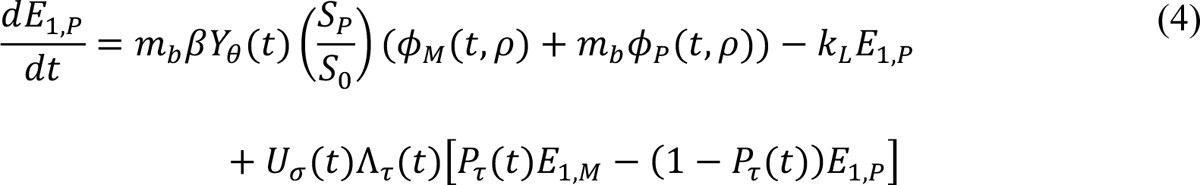

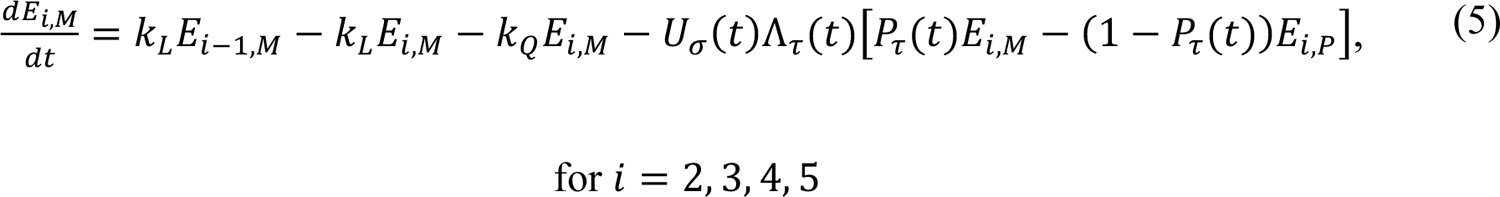

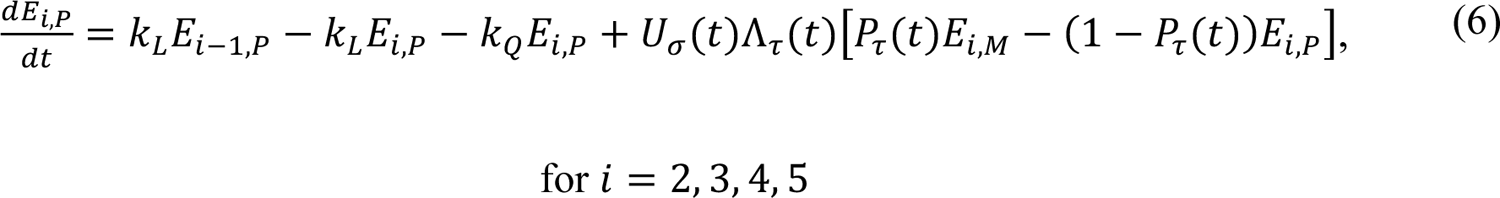

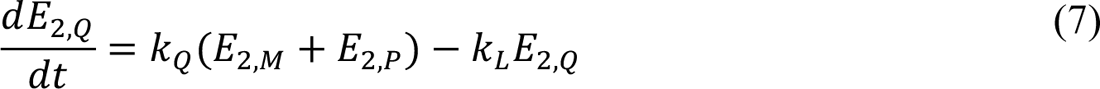

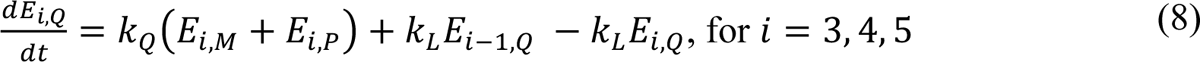

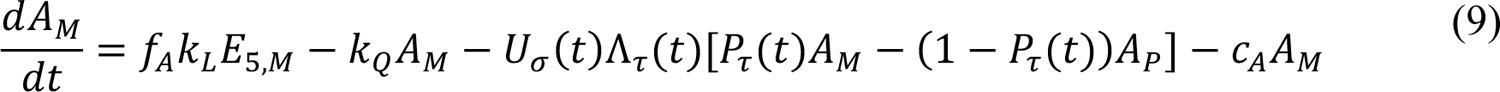

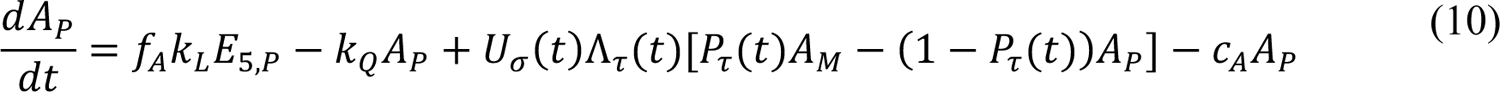

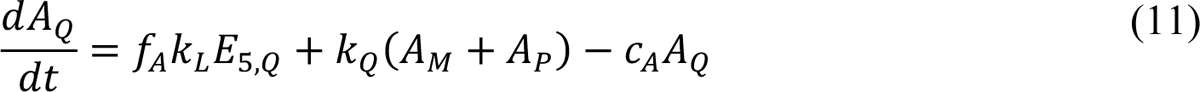

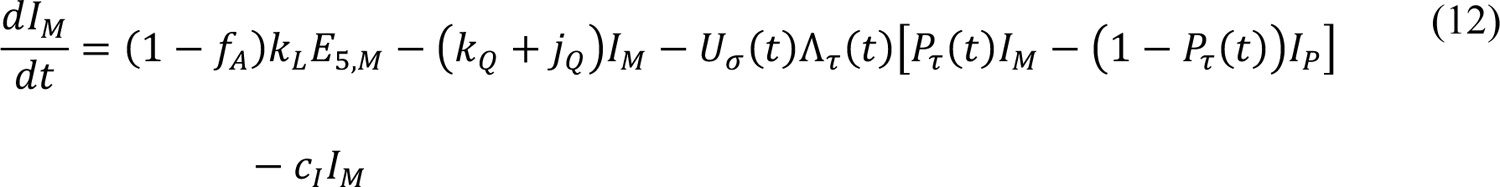

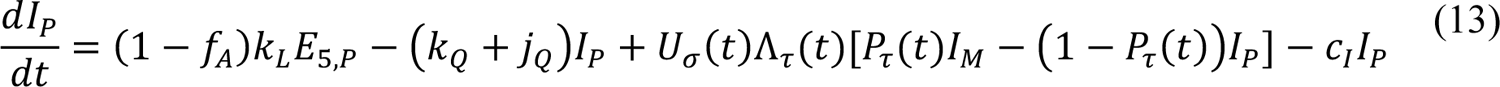

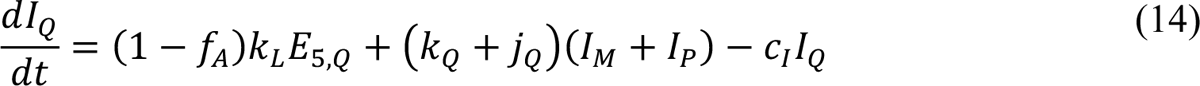

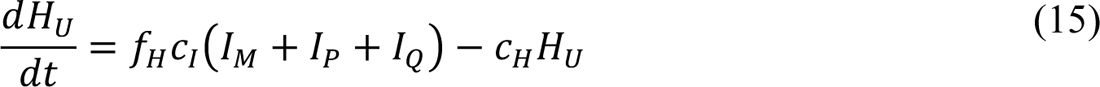

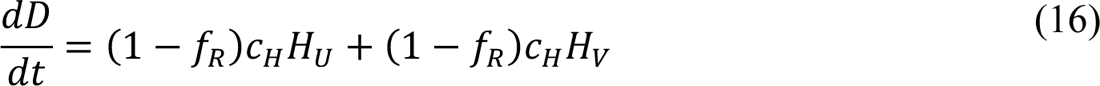

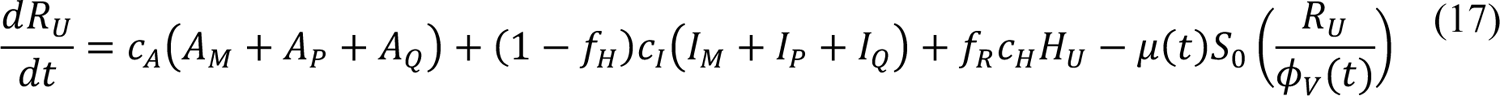

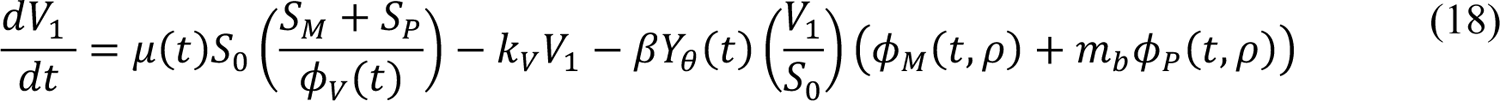

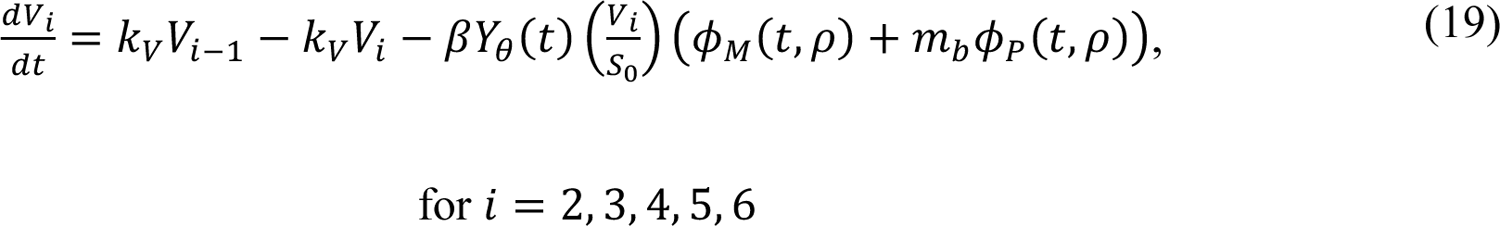

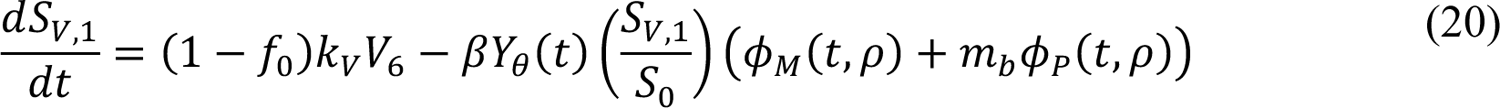

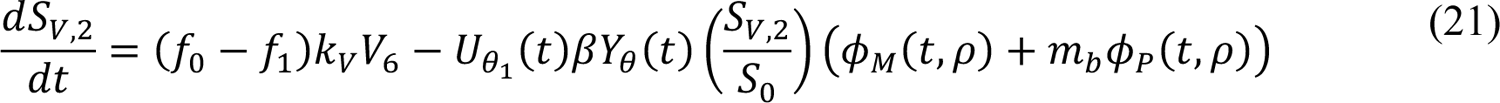

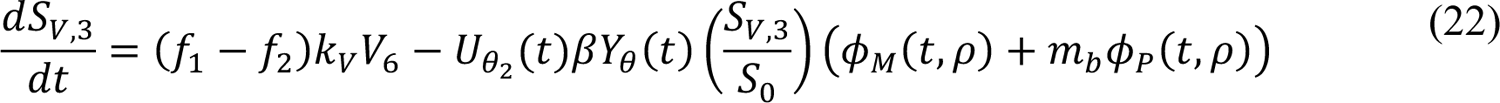

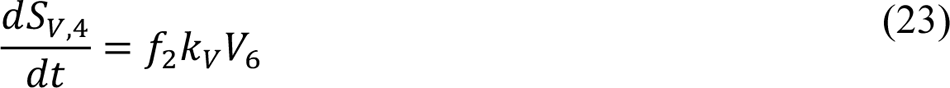

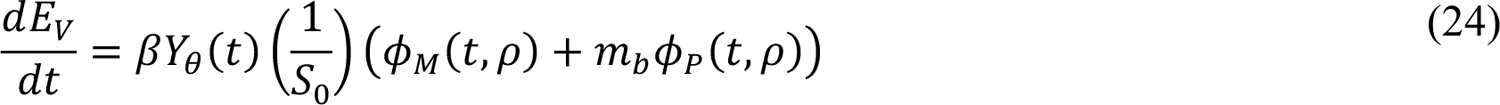

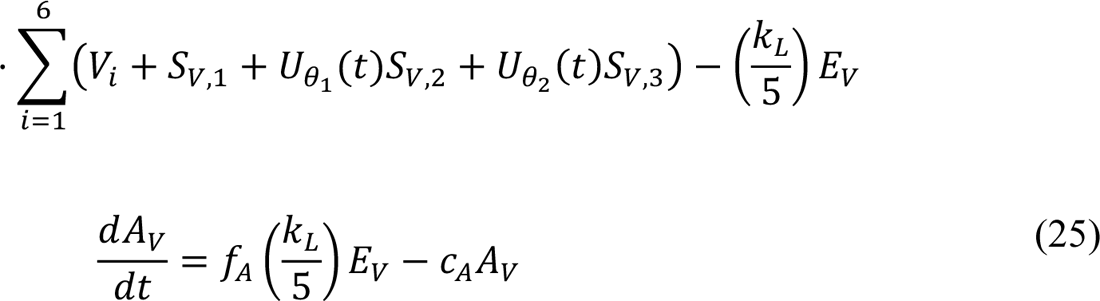

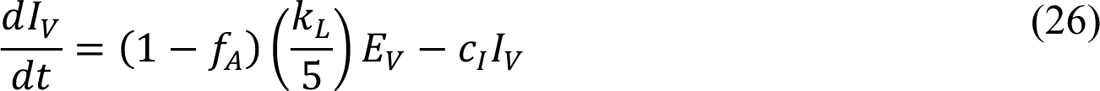

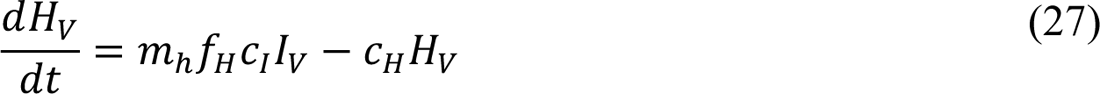

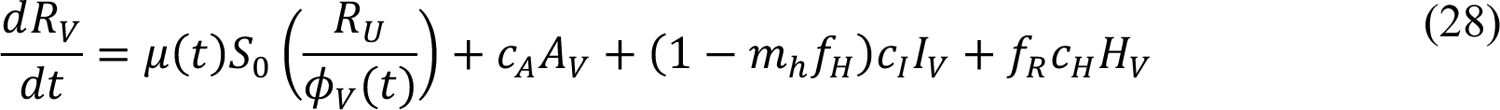

In these equations, the independent variable is time *t*, and the state variables (*S*_*M*_, *S*_*P*_, *E*_1,*M*_, …, *E*_=,*M*_, *E*_1,*P*_, …, *E*_=,*P*_, *E*_),:_, …, *E*_=,:_, *A*_*M*_, *A*_*P*_, *A*_Q_, *I*_*M*_, *I*_*P*_, *I*_Q_, *H*_*U*_, *D*, *R*_*U*_, *V*_1_, …, *V*_&_, *S*_*V*,1_, …, *S*_*V*,(_, *E*_*V*_, *A*_*V*_, *I*_*V*_, *H*_*V*_, and *R*_*V*_) represent 40 (sub)populations (Appendix Table 1), which change over time. Thus, each ODE in Equations (1)–(28) defines the time-rate of change of a population, i.e., the time-rate of change of a state variable. Note that Equations (5), (6), (8) and (19) define 4, 4, 3, and 5 ODEs of the model, respectively. The model is formulated such that *S*_0_, the total population, is a constant. Thus, the model does not account for birth, death for reasons other than COVID-19, immigration, or emigration.

The initial condition associated with Equations (1)–(28) is taken to be *S*_*M*_(*t*_0_) = *S*_1_, *I*_*M*_(*t*_0_) = *I*_1_ = 1, and all other populations (*S*_*P*_, *E*_1,*M*_, …, *E*_=,*M*_, *E*_1,*P*_, …, *E*_=,*P*_, *E*_),:_, …, *E*_=,:_, *A*_*M*_, *A*_*P*_, *A*_Q_, *I*_*P*_, *I*_Q_, *H*_*U*_, *D*, *R*_*U*_, *V*_1_, …, *V*_&_, *S*_*V*,1_, …, *S*_*V*,(_, *E*_*V*_, *A*_*V*_, *I*_*V*_, *H*_*V*_, and *R*_*V*_) are equal to 0. Recall that the parameter *S*_0_ denotes the total region-specific population size. Thus, we assume that the entire population is susceptible at the start of the local epidemic at time *t* = *t*_0_ > 0, where time *t* = 0 corresponds to 00:00 hours (midnight) on January 21, 2020. The parameter *I*_0_ denotes the number of infectious symptomatic persons at the start of the regional epidemic.

In the model, the parameters β, *k*_*L*_, *k*_Q_, *j*_Q_, τ_*A*_, τ_*I*_, τ_*H*_, and *k*_*V*_ are positive-valued rate constants (all with units of d^-1^), and the parameters *m*_*b*_, *m*_2_, *f*_*A*_, *f*_*H*_, *f*_*R*_, *f*_0_ ≥ *f*_1_, *f*_1_ ≥ *f*), and *f*) are (dimensionless) fractions. Brief definitions of parameters are given in Tables 1 and 2.

In the model, the quantities φ_*M*_(*t*, ρ), φ_*P*_(*t*, ρ), and φ_*V*_(*t*) are functions of (time-dependent) state variables (as defined below), which represent the population of infectious persons who are mixing freely (i.e., not practicing social-distancing), the population of infectious persons who are practicing social-distancing (i.e., adopting disease-avoiding behaviors), and the population of persons eligible for vaccination, respectively. The quantities φ_*M*_(*t*, ρ) and φ_*P*_(*t*, ρ) are also functions of ρ ≡ (ρ_*E*_, ρ_*A*_), where ρ_*E*_ (ρ_*A*_) is a dimensionless ratio representing the infectiousness of persons in the incubation phase of infection (the infectiousness of asymptomatic persons in the immune clearance phase of infection) relative to the infectiousness of symptomatic persons with the same social-distancing behavior. The quantity φ_*V*_(*t*) represents the population of persons eligible for vaccination.

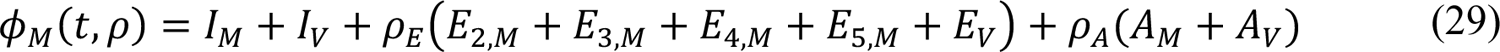

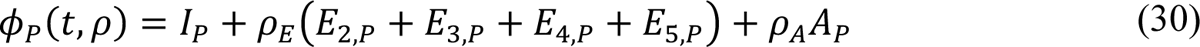

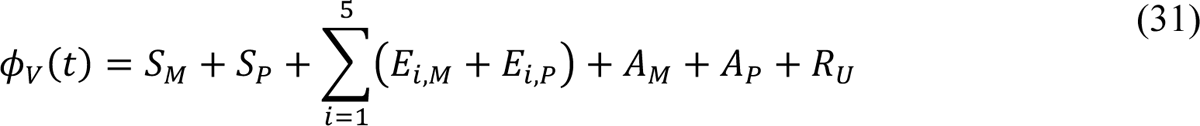

The state variables that appear in these equations represent time-varying populations. Recall that state variables are defined in Appendix Table 1.

In the model, the quantities *U*_σ_(*t*), *U*_1_(*t*), and *U*_2_(*t*) are unit step functions. The values of these functions change from 0 to 1 at the times indicated by the subscripts: σ, the onset time of the initial social-distancing period; θ_1_, the takeoff time of SARS-CoV-2 variant Alpha; and θ), the takeoff time of SARS-CoV-2 variant Delta.

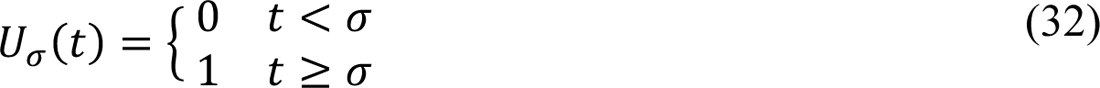

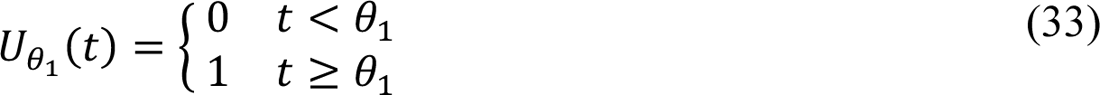

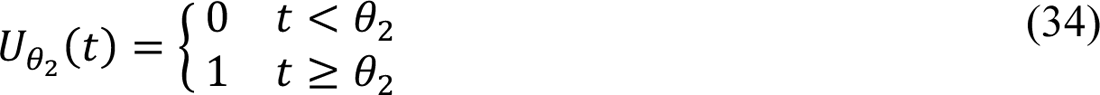

As indicated in Appendix Figure 1, transitions from *S*_*M*_ to *S*_*P*_, for example, become possible at time *t* = σ, transitions from *S*_*V*,)_ to *E*_*V*_ become possible at time *t* = θ_1_, and transitions from *S*_*V*,*_ to *E*_*V*_ become possible at time *t* = θ).

In the model, the quantities *P*_τ_(*t*), and Λ_τ_(*t*) are step functions that characterize changes in social-distancing. The value of *P*_τ_(*t*) determines a setpoint steady-state fraction of susceptible persons who are practicing social-distancing. The value of Λ_τ_(*t*) determines a time scale for approach to the setpoint steady state. Changes in the values of *P*_τ_(*t*) and Λ_τ_(*t*) occur coordinately. These changes occur at times *t* = σ, τ_1_, …, τ_*n*_, where *n* is the number of distinct social-distancing periods beyond an initial social-distancing period. Initially, we took *n* = 7 (i.e., 8 total social-distancing stages). The value of *n* is decremented by 1 (at an inferred time) if *n* ← *n* − 1 is determined to be admissible by a model-selection procedure, which is described below. It should be noted that *p*_0_, *p*_1_, …, *p*_*n*_ are parameters of *P*_τ_(*t*) and that λ_1_, λ_1_, …, λ_*n*_ are parameters of Λ_τ_(*t*). These parameters determine the non-zero values of the step functions over different periods. For example, *p*_1_ is the value of *P*_τ_(*t*) and λ_1_ is the value of Λ_τ_(*t*) for the period *t* ∈.

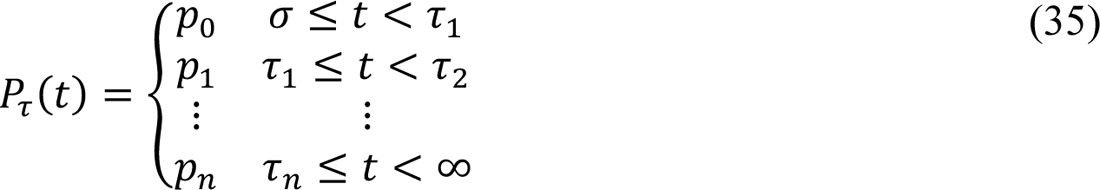

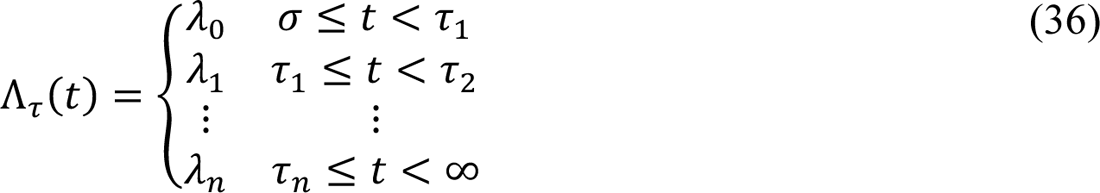

The values of *P*_τ_(*t*) and Λ_τ_(*t*) are 0 for *t* < σ.

In the model, the quantity μ(*t*) is a piecewise linear interpolant to a function μG(*t*) that characterizes the current rate of vaccination. The value of μG(*t*) is determined by the empirical daily rate of vaccination, and thus, can vary from day to day. Daily vaccination data were extracted from the Covid Act Now database and the *Democrat and Chronicle* newspaper [2] using the Covid Act Now Data ApI [1] and web scraping. We will use μ_+_ to refer to the value of μG(*t*) for *t* ∈ [*t*_+_, *t*_+31_), where time *t*_+_ corresponds to midnight on the *i*th day after January 21, 2020.

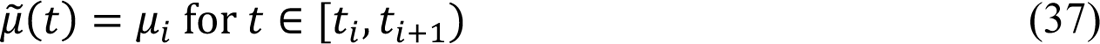

Settings for μ_+_ were made such that μ_+_*S*_0_ × 1 d is the number of vaccinations completed in the nearest past 1-d period according to Covid Act Now data and the *Democrat and Chronicle* newspaper.

In the model, the quantity *Y*(*t*) is a step function that quantifies how disease transmissibility increases upon emergence of SARS-CoV-2 variants Alpha and Delta. Initially, *Y*(*t*) = 1. The value of *Y*(*t*) is increased (by an inferred factor greater than 1 at an inferred time, θ_1_ or θ)) if the change is determined to be admissible by a model-selection procedure, which is described below. It should be noted that *y*_1_ and *y*) are parameters of *Y*(*t*). These parameters determine the values of the step function *Y*(*t*) over different periods: *y*_1_ is the value of *Y*(*t*) for the period *t* ∈ [θ_1_, θ)) and *y*) is the value of *Y*(*t*) for the remaining period of concern (with Delta as the dominant circulating viral strain).

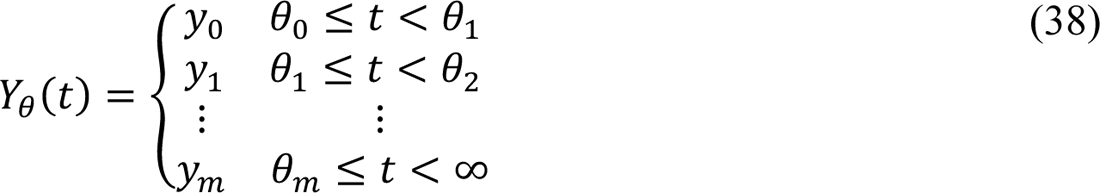

where *m* is the number of viral variants that have emerged up to the current time, θ_0_ ≡ *t*_1_, and *y*_0_ ≡ 1. Here, we consider *m* = 2.

### Equations of the Auxiliary Measurement Model

As in the study of Lin et al. [3], we assumed that only symptomatic persons are detected in testing. The accumulation of symptomatic persons is governed by

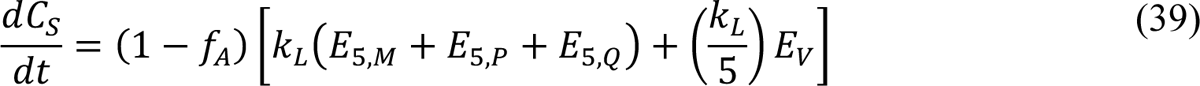

where *C*_*S*_(*t*) is the cumulative number of symptomatic persons (cases) at time *t*. Here, unlike in the study of Lin et al. [3], the expression for *C*_*S*_(*t*) accounts for exposed persons in quarantine. Initially, *C*_*S*_ = 0. We numerically integrated Appendix Equation (39) together with the ODEs of the compartmental model. From the trajectory for *C*_*S*_, we derive a prediction for the expected number of new COVID-19 cases reported on calendar date *D*_i_, *I*(*t*_+_, *t*_+31_), using the following equation:

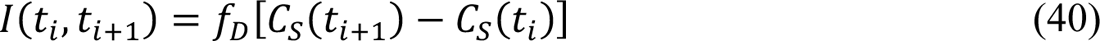

where *f*_*D*_is an adjustable region-specific parameter characterizing the time-averaged fraction of symptomatic cases detected and reported, *t*_+_ corresponds to midnight on the *i*th day after January 21, 2020, and *t*_+31_ − *t*_+_ is the reporting interval (1 d). We compare *I*(*t*_+_, *t*_+31_) to δ*C*_+_, the number of new cases reported on calendar date *D*_i_.

### Definition of the Likelihood Function

Bayesian inference relies on the definition of a likelihood, which here serves the purpose of assessing the compatibility of available surveillance data with adjustable (free) parameter values. Let us use {δ*C*_+_}^d^_i=0_ to denote the daily case reporting data available between 0 and *d* days after midnight on January 21, 2020 (the date of the first case report in the US) and let *D* = {δ*C*_+_}^*d*^. Let us use θ_*C*_(*n*, *m*) to denote the set of adjustable (free) parameter values. The number of adjustable parameters, |θ_*C*_|, depends on *n*, the number of social-distancing periods considered beyond an initial social-distancing period, and *m*, the number of SARS-CoV-2 variants under consideration. As in the study of Lin et al. [3], we assume that δ*C*_+_, the number of new COVID-19 cases detected over a 1-d period and reported on calendar date *D*_i_ for a given region, is a random variable and its expected value follows a model-derived deterministic trajectory given by *I*(*t*_+_, *t*_+31_) (Equation 40). We assume that day-to-day fluctuations in the random variable are independent and characterized by a negative binomial distribution NB(τ, *q*_+_), which has two parameters, τ > 0 and *q*_+_ ∈ (0,1). Note that D[NB(τ, *q*_+_)] = τ(1 − *q*_+_)/*q*_+_. We assume that this distribution has the same dispersion parameter τ across all case reports. With these assumptions, we arrive at the following likelihood function:

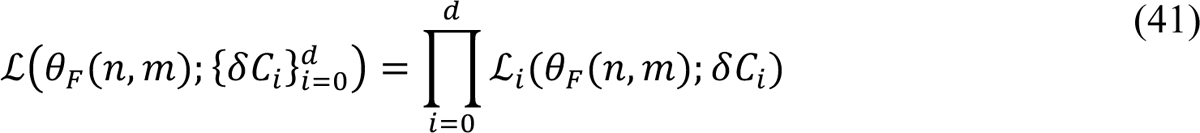

where

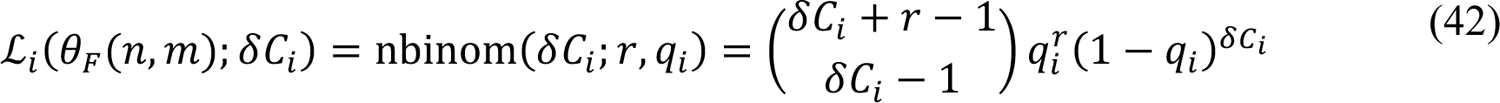

and

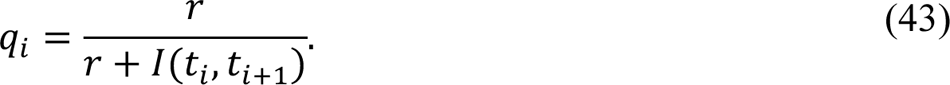

In these equations, *i* is an integer indicating the date *D*_+_ and period (*t*_+_, *t*_+31_); nbinom(δ*C*_+_; τ, *q*_+_) is the probability mass function of the negative binomial distribution NB(τ, *q*_+_), and θ_*C*_(*n*, *m*) = {*t*_0_, β, σ, τ_1_, …, τ_*n*_, *p*_0_, *p*_1,_ …, *p*_*n*_, λ_0_, λ_1_, …, λ_*n*_, θ_1_, …, θ_*m*_, *y*_1_, …, *y*_*m*_, *f*_*D*_, τ} for *n* ≥ 1 and *m* ≥ 1; θ_*C*_(0,0) = {*t*_0_, β, *p*_0_, λ_1_, *f*_*D*_, τ}.

### parameters

Each model parameter is briefly described in Tables 1–3. These parameters have either fixed values or adjustable values (i.e., values inferred from surveillance data). The fixed values may be universal (i.e., applicable to all MSAs of interest) or MSA-specific. All inferred parameter values are MSA-specific. In addition, the measurement model (Appendix Equations 39 and 40) has one adjustable MSA-specific parameter, *f*_*D*_, and the likelihood function (Appendix Equations 41–43) has one adjustable MSA-specific parameter, τ. Values of the other likelihood parameters, *q*_0_, …, *q*_*d*_, are constrained and are determined using Appendix Equation 43.

### Original model of Lin et al.

The model shares 19 + 3*n* parameters with the model of Lin et al. [3], including parameters that define the initial condition (*t*_0_, *I*_1_, and *S*_0_). (Recall that *n* is the number of social-distancing periods being considered beyond the initial social-distancing period.) The shared parameters are *t*_0_, *I*_1_, *S*_2_, β, σ, τ_1_, …, τ_*n*_, *p*_0_, …, *p*_*n*_, λ_0_, …, λ_*n*_, ρ_*A*_, ρ_*E*_, *m*_*b*_, *f*_*A*_, *f*_*H*_, *f*_*R*_, *k*_*L*_, *k*_Q_, *j*_Q_, τ_*A*_, τ_*H*_, and τ_*I*_. As in the study of Lin et al. [3], we inferred MSA-specific values for the following parameters: *t*_0_, β, σ, *p*_1_, …, *p*_*n*_, and λ_1_, …, λ_*n*_. We also inferred MSA-specific values for τ_1_, …, τ_*n*_ provided that *n* ≥ 1. As in the study of Lin et al. [3], the remaining 14 parameters shared between the old and new models (*I*_0_, *S*_1_, ρ_*A*_, ρ_*E*_, *m*_*b*_, *f*_*A*_, *f*_*H*_, *f*_*R*_, *k*_*L*_, *k*_Q_, *j*_Q_, τ_*A*_, τ_*H*_, and τ_*I*_) were taken to have fixed values, and we adopted the settings of Lin et al. [3] for these parameters (Table 3). These settings are universal except for the setting for *S*_1_, the total population, which is MSA-specific.

### Extension of the model of Lin et al.

Our extension of the model of Lin et al. [3] introduces 5 + 2(*m* + 1) + (*d* + 1) parameters, where *m* (= 0, 1 or 2) is the number of SARS-CoV-2 variants being considered and *d* is the number of days since January 21, 2020: θ_1_, …, θ_*m*_, *y*_0_, …, *y*_*m*_, *m*_2_, *f*_0_, *f*_1_, *f*), *k*_*V*_, and μ_0_, …, μ_*d*_. The θ and *y* parameters are variant takeover times and transmissibility factors, respectively, except that the value of θ_1_ is defined as *t*_0_ and the value of *y*_0_ is defined as 1. The Alpha transmissibility factor *y*_1_, the Alpha takeoff time θ_1_, the Delta transmissibility factor *y*), and the Delta takeoff time θ) were inferred for each MSA with *m* = 2 (cf. Table 1). The transmissibility factors were each constrained to be greater than or equal to 1. The settings for μ_1_, …, μ_*d*_ are empirical and MSA-specific. Each μ_+_ is set such that μ_+_*S*_0_ × 1 d is the number of vaccinations completed over the past 1-d period nearest to the *i*th day after January 21, 2020. As noted earlier, the number of completed vaccinations was obtained for each MSA from Covid Act Now and the *Democrat and Chronicle* newspaper [2] using the Covid Act Now Data ApI [1] and web scraping. In the spreadsheet accessed daily from Covid Act Now, the ‘metrics.vaccinationsCompletedRatio’ column gives the percentage of the total population that has received the recommended number of doses: one dose for Ad26.CoV2.S (Janssen, Johnson & Johnson) or two doses for mRNA-1273 (Moderna) and BNT162b2 (pfizer-BioNTech). As a simplification, we considered all completed vaccinations to be equivalent. The parameters *m*_2_, *f*_0_, *f*_1_, *f*), and *k*_*V*_ were assigned fixed universal estimates (Table 3). Each of these estimates is explained below.

### Estimation of *k*_*V*_

The rate constant *k*_*V*_ characterizes the rate of transition out of compartment *V*_+_ for *i* = 1, …, *n*_*V*_. Recall that, in the model, susceptible persons enter *V*_1_ upon vaccination (Figure 1, Appendix Figure 1). The values of *n*_*V*_ (= 6) and *k*_*V*_ (= 0.3 d^-1^) were selected so that the time a person spends in *V*_1_, …, *V*_*n*)_, which we will denote as *t*_*V*_, is distributed approximately the same as, the waiting time between vaccination of a previously uninfected person and detection of vaccine-induced SARS-CoV-2-specific IgG antibodies [4] (Appendix Figure 2). According to the model, the time that a person spends in *V*_1_, …, *V*_*n*)_is distributed according to the probability density function *f*(*t*_*V*_; *n*_*V*_, *k*_*V*_) = *k*^*n*)^*t*^*n*)–1^*d*^-*k*)*t*)^/(*n*_*V*_ − 1)!, i.e., *t*_*V*_ is Erlang distributed with shape parameter *n*_*V*_ = 6 and rate parameter *k*_*V*_ = 0.3 d^-1^. As can be seen in Appendix Figure 2, the cumulative distribution function of this Erlang distribution reasonably captures the empirical cumulative distribution of waiting times observed in the longitudinal study of Korodi et al. [4].

Thus, in the model, passage through *V*_1_, …, *V*_&_ with rate constant *k*_*V*_ = 0.3 d^-1^ accounts for the variable and significantly non-zero amount of time required for development of a protective antibody response after vaccination.

### Estimation of *f*_0_, *f*_1_, and *f*_2_

The parameters *f*_0_> *f*_1_, *f*_1_ > *f*), and *f*) are fractions that characterize the average effectiveness of vaccines used in the US and that determine the sizes of (mutually exclusive) subpopulations of vaccinated persons having different susceptibilities to productive infection (i.e., an infection that can be transmitted to others): *S*_*V*,1_, *S*_*V*,)_, *S*_*V*,*_, and *S*_*V*,(_ (Figure 1, Appendix Figure 1). We assume that persons in the *S*_*V*,1_ subpopulation are susceptible to productive infection by any of the viral strains considered, and in contrast, we assume that persons in the *S*_*V*,(_ subpopulation are susceptible to productive infection by none of the viral strains considered. persons in the *S*_*V*,)_ subpopulation are taken to be susceptible to productive infection by the Alpha and Delta variants but not viral strains in circulation before the emergence of Alpha. persons in the *S*_*V*,*_ subpopulation are taken to be susceptible to productive infection by the Delta variant but not the Alpha variant or viral strains in circulation before the emergence of Alpha. The quantity 1 − *f*_0_ defines the fraction of vaccinated persons who enter the *S*_*V*,1_subpopulation after exiting *V*_&_, the quantity *f*_0_ − *f*_1_ defines the fraction of vaccinated persons who enter the *S*_*V*,)_ subpopulation after exiting *V*_&_, the quantity *f*_1_ − *f*) defines the fraction of vaccinated persons who enter the *S*_*V*,*_ subpopulation after exiting *V*_&_, and *f*) defines the fraction of vaccinated persons who enter the *S*_*V*,_ (subpopulation after exiting *V*_&_. We take *f*_0_ to characterize vaccine effectiveness before the emergence of Alpha. According to Thompson et al. [5], vaccine effectiveness was initially 90%. Thus, we set *f*_1_ = 0.9. We take *f*_1_ to characterize vaccine effectiveness after the emergence of Alpha but before the emergence of Delta. According to puranik et al. [6], in May 2021, vaccine effectiveness was 81%. Thus, we set *f*_1_ = 0.81. We take *f*) to characterize vaccine effectiveness after the emergence of Delta. According to Tang et al. [7], the effectiveness of two doses of the pfizer-BioNTech vaccine (BNT162b2) against Delta is 53.5% and the effectiveness of two doses of the Moderna vaccine (mRNA-1273) against Delta is 84.8%. Taking the average of these figures, we set *f*) = 0.69.

### Estimation of *m*_*h*_

The parameter *m*_2_ characterizes the reduced risk of severe disease for a vaccinated person in the case of a breakthrough infection. We set *m*_2_ = 0.04, i.e., we assumed that there is a 25-fold reduction in the risk of severe disease for infected persons who have been vaccinated, which is consistent with the observations of Lopez Bernal et al. [8].

### Notable New Modeling Assumptions

It should be noted that we treat the incubation period for newly infected (exposed) vaccinated persons differently than for newly infected (exposed) unvaccinated persons (Figure 1, Appendix Figure 1). For unvaccinated persons, as in the study of Lin et al. [3], we divide exposed persons in the incubation period of infection into five subpopulations: *E*_1,*M*_, …, *E*_=,*M*_for exposed persons who are mixing (i.e., persons who are not practicing social-distancing), *E*_1,*P*_, …, *E*_=,*P*_ for exposed persons who are practicing social-distancing, and *E*_1,:_, …, *E*_=,:_ for exposed quarantined persons. persons move through the five stages of the incubation period sequentially. In contrast, as a simplification, for vaccinated persons, we consider only a single exposed population: *E*_*V*_. We take persons to exit *E*_*V*_ with rate constant *k*_*L*_/5 (Appendix Figure 1). With this choice, the duration of the incubation period of infection is the same, on average, for both vaccinated and unvaccinated persons. The average duration is 5/*k*_*L*_ (about 5 d) in both cases. The difference is that the duration of the incubation period is Erlang distributed for unvaccinated persons, as discussed by Lin et al. [3], but exponentially distributed for vaccinated persons.

As indicated in Equation (29), we take vaccinated persons with productive infections to be equally as infectious as unvaccinated persons.

As noted earlier, we take all vaccinated persons to be mixing (i.e., to not be practicing social-distancing). Thus, populations of infected vaccinated persons (*E*_*V*_, *I*_*V*_, and *A*_*V*_) contribute to φ_*M*_(*t*) (Appendix Equation 29) but not φ_*P*_(*t*) (Appendix Equation 30).

As indicated in Appendix Equation 31, we consider pre-symptomatic exposed and asymptomatic unvaccinated persons to be eligible for vaccination and, thus, these persons contribute to the consumption of vaccine doses (i.e., these persons account for a portion of the number of completed vaccinations on a given day *i*, μ_+_*S*_0_ × 1 d). However, we do not move these persons to vaccinated compartments. The reason is that exposed and asymptomatic persons are expected to develop immunity faster through recovery from infection (i.e., movement to *R*_*U*_) than from vaccination.

As indicated in Appendix Equation 31, we do not consider symptomatic, quarantined, severely ill and hospitalized/isolated-at-home, and deceased persons to be eligible for vaccination.

### Inference Approach

Recall that θ_*C*_ denotes the set of all adjustable parameters. As in the study of Lin et al. [3], for each MSA, we inferred MSA-specific adjustable parameter values θ_*C*_ using all MSA-specific surveillance data available up to a specified day of inference *D*_R_(i.e., the *d*th day after January 21, 2020). We took a Bayesian inference approach, meaning that, for a given dataset, we generated parameter posterior samples (a collection of θ_*C*_’s) through Markov chain Monte Carlo (MCMC) sampling. The parameter posterior samples provide a probabilistic characterization of the adjustable parameter values consistent with the dataset used in inference. By drawing samples from the parametric posterior distribution, we generated a posterior predictive distribution for *I*(*t*_+_, *t*_+31_) for each *i* of interest. We considered all days from January 21, 2020 to October 31, 2021. In other words, for each *i* of interest, a prediction for *I*(*t*_+_, *t*_+31_) was generated for each of many θ_*C*_’s drawn randomly (with uniform probability) from the parametric posterior distribution. The resulting distribution of *I*(*t*_+_, *t*_+31_) values is the posterior predictive distribution for *I*(*t*_+_, *t*_+31_). Recall that *I*(*t*_+_, *t*_+31_) is given by Appendix Equation 40 and corresponds to D[δ*C*_+_], the expected number of new COVID-19 cases detected over a 1-d surveillance interval and reported for the *i*th day after January 21, 2020. Observation noise was injected into the posterior predictive distributions by replacing each sampled value for *I*(*t*_+_, *t*_+31_) with *X*_i_∼NB(τ, *q*_+_), where τ is a member of the sampled set of parameter values θ_*C*_ used to generate the prediction of *I*(*t*_+_, *t*_+31_) and *q*_+_ is given by Equation 43.

According to Bayes’ theorem, given surveillance data *D* = {δ*C*_+_}^*d*^, the parametric posterior is given as

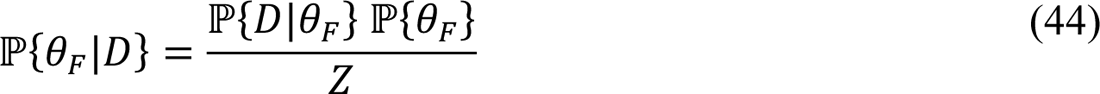

where ℙ{θ_*C*_} is the prior (which is formulated to capture knowledge of θ_*C*_ external to *D* or to express lack of such knowledge), ℙ{*D*|θ_*C*_} is the likelihood defined by Appendix Equations 41– 43, and *Z* is a normalizing constant. We assumed a proper uniform prior, i.e., for each adjustable parameter, we assumed that all values between specified lower and upper bounds are equally likely before consideration of *D*. We used the same bounds as in the study of Lin et al. [3]. Then, we used an adaptive MCMC (aMCMC) algorithm [9] to generate samples from ℙ{*D*|θ_*C*_} ℙ{θ_*C*_}, which is proportional to ℙ{θ_*C*_|*D*}. Thus, the relative probabilities of parameter sets θ_*C*_ according to ℙ{θ_*C*_|*D*} are correctly represented by the samples.

Specifically, the adaptive MCMC algorithm [9] generates samples from the multivariate parametric posterior for adjustable model parameters (*t*_0_, β, and parameters for variant emergence and social-distancing), the measurement model parameter *f*_*D*_, and the likelihood parameter τ (Tables 1 and 2). This algorithm is available within the pyBioNetFit software package [10]. Use of the algorithm was performed as described by Lin et al. [3]. The report of Neumann et al. [10] includes helpful general usage advice, which was followed in this study.

Inference jobs were executed on a computer cluster. For each inference job, a total of 25 chains were generated, and the chain with the best mixing and convergence properties was selected for subsequent analyses.

Each inference was conditioned on the compartmental model of Figure 1 (Appendix Equations 1–38), settings for the structural parameters *m* (the number of SARS-CoV-2 variants under consideration) and *n* (the number of social-distancing periods under consideration beyond an initial social-distancing period), the measurement model (Appendix Equations 39 and 40), and fixed parameter estimates (Tables 1 and 2), including empirical daily *per capita* vaccination rates (i.e., the settings for μ_+_ in Appendix Equation 37). We assumed a proper uniform prior for each adjustable parameter [3] and a negative binomial likelihood function (Appendix Equations 41– 43). Use of proper uniform priors means that MAp estimates are maximum likelihood estimates (MLEs). In each inference, the data entering the likelihood function, *D* = {δ*C*_+_}^*d*^ (Appendix Equation 41), were MSA-specific daily reports of newly detected COVID-19 cases available up to the date of inference *D*_R_ (i.e., the *d*-th day after January 21, 2020). Thus, all inferences are region-specific and time-dependent.

### Use of Model Selection to Determine Intervals of Step Functions

Variant takeover times, θ = (θ_1_, θ)), and start times of social-distancing periods, τ = (σ, τ_1_, …, τ_*n*_), were inferred from data; however, changes of the associated time-dependent step functions, *Y*(*t*), *P*_τ_(*t*), and Λ_τ_(*t*), were introduced only when an increase in model complexity was deemed to be justified. Each decision to introduce variant takeover or start of a new social-distancing period (beyond the initial period) was made using a model-selection procedure, which is described below. It should be noted that *y*_1_ and *y*) are parameters of *Y*(*t*), *p*_0_, *p*_1_, …, *p*_*n*_ are parameters of *P*_τ_(*t*), and λ_1_, λ_1_, …, λ_*n*_are parameters of Λ_τ_(*t*). These parameters determine the values of the step functions over different periods. For example, for *n* ≥ 1, *p*_1_ is the value of *P*_τ_(*t*) and λ_1_ is the value of Λ_τ_(*t*) for the period *t* ∈ [τ_1_, τ)). Similarly, *y*_1_ is the value of *Y*(*t*) for the period *t* ∈ [θ_1_, θ)), and *y*) is the value of *Y*(*t*) for the period *t* ∈ [θ), *t*_final_), where *t*_final_ corresponds to the date of inference (October 31, 2021).

We started with a setting of *n* = 7 for each MSA of interest (i.e., 8 total social-distancing stages). To determine if *n* could be reduced, we conducted parsimony checks. In a parsimony check, 100 MLE curves, each constituting a fit to the data, were generated via optimization jobs. In these jobs, the total number of social-distancing stages in the model (*n* + 1) was set at 1 less than for the current proposed best fit. Each of the 100 fits was visually inspected to determine whether or not the following criteria held: 1) the quality of fit is acceptable (i.e., comparable to what is obtained with the current proposed best fit); 2) Alpha and Delta surges (identified by sequencing data) are explained at least partly by increased transmissibility; 3) social-distancing setpoint parameter values are feasible (i.e., each setpoint parameter takes on a value between 0.2 and 0.8); and 4) social-distancing changes proximal to an Alpha or Delta surge (if any) occur only after an increase in transmissibility. If one or more of these conditions was not satisfied, we accepted the proposed best fit as the most parsimonious fit to the data. If all conditions held, we updated the proposed best fit, and the parsimony check was repeated for a model with one less social-distancing stage. In the Appendix, we show examples of violations of parsimony-check criteria for the MSA surrounding Houston (Appendix Figure 3).

### Simulations

After specification of parameter values (Tables 1–3), we used the Scipy (https://www.scipy.org) interface to LSODA [12] to numerically integrate the system of coupled ODEs consisting of the 40 ODEs of the compartmental model and the 1 ODE of the measurement model (Appendix Equations 1–39). The initial condition was defined by settings for *t*_0_, *I*_1_, and *S*_2_ (Tables 1–3). Integration combined with use of Appendix Equation 40 yielded a prediction of the expected number of new cases detected for each 1-d surveillance period of interest in the past or future: *I*(*t*_+_, *t*_+31_), where *t*_+_ corresponds to midnight on the *i*th day after January 21, 2020. To account for randomness in case detection and reporting, we replaced *I*(*t*_+_, *t*_+31_) with *X*_+_∼NB(τ, *q*_+_), where *q*_+_ is given by Appendix Equation 43.

---

**Appendix Table 1.** State variables of the compartmental model.

| State variable<br>(population) | Description |
| --- | --- |
| $S_M$ | Population of susceptible unvaccinated persons who are mixing (i.e., not practicing social-distancing) |
| $S_P$ | Population of susceptible unvaccinated persons who are practicing social-distancing |
| $S_{V,1}$ | Population of vaccinated unexposed persons who developed an immune response to vaccination that does not protect against productive infection by ancestral strains or the Alpha and Delta variants |
| $S_{V,2}$ | Population of vaccinated unexposed persons who developed an immune response to vaccination that protects against productive infection by ancestral strains but not the Alpha or Delta variants |
| $S_{V,3}$ | Population of vaccinated unexposed persons who developed an immune response to vaccination that protects against productive infection by ancestral strains and the Alpha variant but not the Delta variant |
| $S_{V,4}$ | Population of vaccinated unexposed persons who developed an immune response to vaccination that protects against productive infection by ancestral strains and the Alpha and Delta variants |
| $V_i$ ($i = 1, \dots, 6$) | Population of vaccinated persons in the $i$th stage of immune response to vaccination |
| $E_{i,M}$ ($i = 1, \dots, 5$) | Population of exposed unvaccinated persons in the $i$th stage of the incubation period of infection and who are mixing |
| $E_{i,P}$ ($i = 1, \dots, 5$) | Population of exposed unvaccinated persons in the $i$th stage of the incubation period of infection and who are practicing social-distancing |
| $E_{i,Q}$ ($i = 2, \dots, 5$) | Population of exposed unvaccinated persons in the $i$th stage of the incubation period of infection and who are quarantined |
| $E_V$ | Population of vaccinated persons in the incubation period of a productive infection (i.e., an infection that can be transmitted to others) |
| $A_M$ | Population of asymptomatic unvaccinated persons who are in the immune clearance phase of infection and who are mixing |
| $A_P$ | Population of asymptomatic unvaccinated persons who are in the immune clearance phase of infection and who are practicing social-distancing |
| $A_Q$ | Population of asymptomatic unvaccinated persons who are in the immune clearance phase of infection and who are quarantined |
| $A_V$ | Population of asymptomatic vaccinated persons who are in the immune clearance phase of a productive infection (i.e., an infection that can be transmitted to others) |
| $I_M$ | Population of infectious, symptomatic, and unvaccinated persons with mild disease who are mixing |
| $I_P$ | Population of infectious, symptomatic, non-vaccinated, and infectious persons with mild disease who are practicing social-distancing |
| $I_Q$ | Population of infectious, symptomatic, and unvaccinated persons with mild disease who are quarantined |
| $I_V$ | Population of infectious, symptomatic, and vaccinated persons with mild disease |
| $R_U$ | Population of recovered unvaccinated persons |
| $R_V$ | Population of recovered vaccinated persons |
| $H_U$ | Population of unvaccinated persons with severe disease who are hospitalized or isolated at home |
| $H_V$ | Population of vaccinated persons with severe disease who are hospitalized or isolated at home |
| $D$ | Population of deceased persons |

**Appendix Figure 1.**
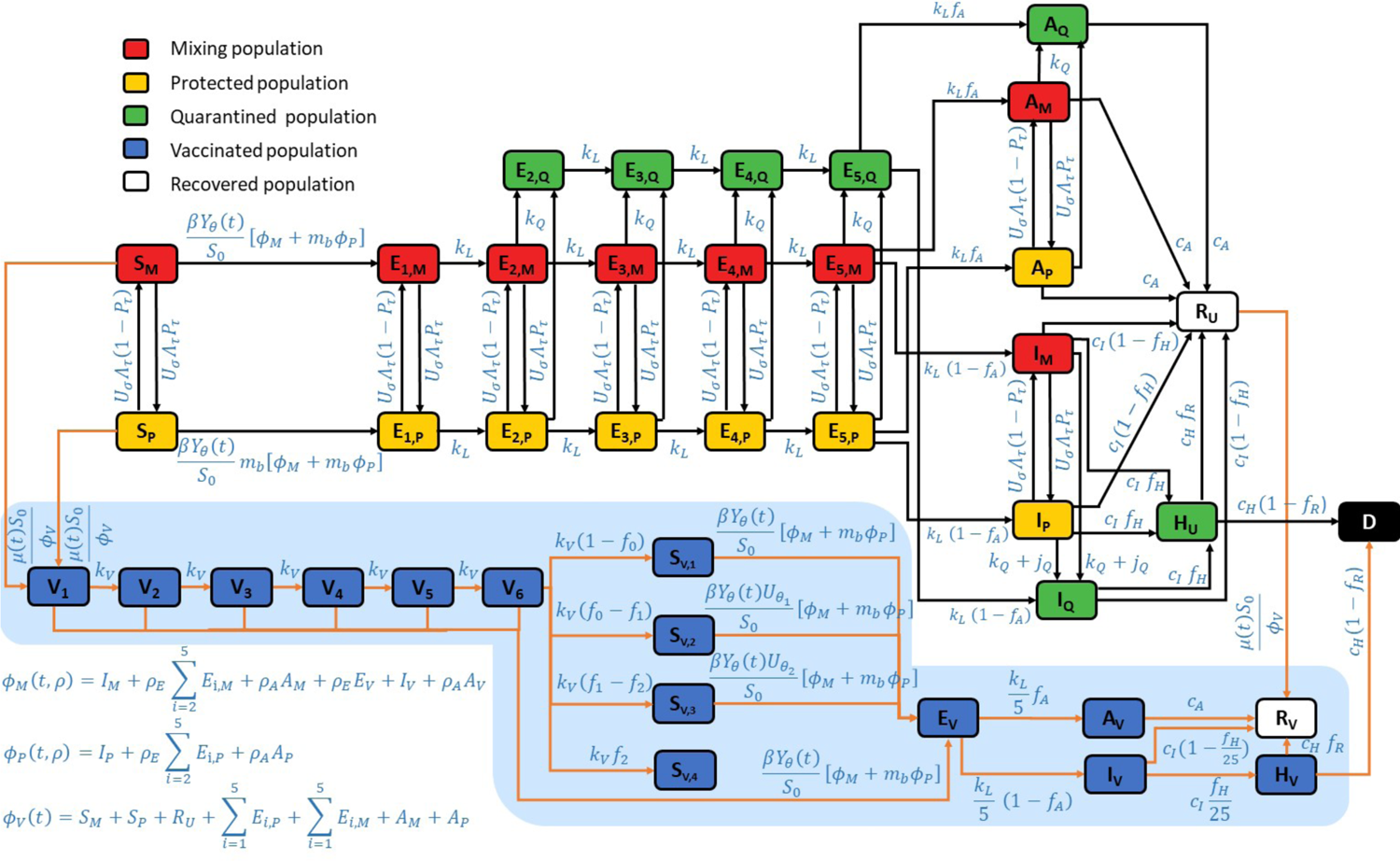
Expanded illustration of the new compartmental model. In the extended model, vaccination was considered by allowing susceptible and recovered persons to transition into a vaccinated compartment, either *V*_1_ and *R*_*V*_. Susceptible (recovered) persons who have completed vaccination move into the *V*_1_(*R*_*V*_) compartment. The susceptible persons who move into *V*_1_are drawn from *S*_*M*_(populated by susceptible persons who are mixing and unprotected by social-distancing) and from *S*_*P*_ (populated by susceptible persons who are protected by social-distancing). After susceptible persons enter *V*_1_, they can move through a series of additional compartments (*V*)through *V*_&_), which are included to capture the time needed for immunity to develop after completion of vaccination. We estimate that the time needed to acquire immunity after vaccination is approximately three weeks based on longitudinal studies of anti-spike protein IgG levels [4]. persons who exit the *V*_&_ compartment without becoming infected enter one of the following compartments: *S*_*V*,1_, *S*_*V*,)_, *S*_*V*,*_, or *S*_*V*,(_. persons in *S*_*V*,1_ are taken to remain susceptible to productive infection by all SARS-CoV-2 strains of interest (Alpha, Delta, and ancestral strains). persons in *S*_*V*,)_ are taken to be susceptible to SARS-CoV-2 Alpha and Delta variants. persons in *S*_*V*,*_ are taken to be susceptible to Delta. persons in *S*_*V*,(_ are taken to be protected against all strains of interest. Infection of persons in *S*_*V*,*_ is only allowed if Delta is present, i.e., at times *t* > θ). Infection of persons in *S*_*V*,)_ is only allowed if Alpha or Delta is present, i.e., at times *t* > θ_1_. Vaccinated persons in compartments *V*_1_ through *V*_&_ and compartment *S*_*V*,1_ are allowed to become infected at any time, at which point they transition to compartment *E*_*V*_, consisting of vaccinated persons who were exposed before development of vaccine-induced immunity. persons in compartment *S*_*V*,)_ are allowed to become infected if *t* ≥ θ_1_. Similarly, persons in compartment *S*_*V*,*_ are allowed to become infected if *t* ≥ θ). possible outcomes for persons in *E*_*V*_are taken to be the same as those for unvaccinated exposed persons; however, the incubation period is taken to be distinct. persons in *E*_*V*_ can experience asymptomatic disease (upon entering *A*_*V*_) or they can become symptomatic (upon entering *I*_*V*_). persons in *A*_*V*_ eventually recover, entering compartment *R*_*V*_. persons in *I*_*V*_ can progress to severe disease (upon entering *H*_*V*_) or recover (upon entering *R*_*V*_). persons in *H*_*V*_ either recover (moving into *R*_*V*_) or die (moving into *D*). persons who have recovered from infection, in the *R*_*U*_ compartment, move directly into the *R*_*V*_ compartment upon vaccination. persons in the *R*_*U*_ and *R*_*V*_ compartments are taken to have full immunity. The vaccination rate at which susceptible and recovered persons move into vaccinated compartments is updated daily for consistency with the empirical overall rate of vaccination, which we extract daily from the COVID Act Now Data ApI [1] and the *Democrat and Chronicle* newspaper [2]. The relative values of the vaccination rate are set such that each person eligible for vaccination has the same probability of being vaccinated. All unvaccinated persons are taken to be eligible for vaccination except symptomatic persons (in compartments *I*_*M*_ and *I*_*P*_), persons who are hospitalized or severely ill at home (in compartment *H*), quarantined persons (in the various compartments labeled with a Q subscript), and deceased persons (in compartment *D*). It should be noted that asymptomatic, non-quarantined persons (in compartments *A*_*M*_ and *A*_*P*_) and presymptomatic, non-quarantined persons (in the *E* compartments) are taken to be eligible (and to influence the vaccination rate constants) but, as a simplification, these persons are not explicitly tracked as vaccinated or unvaccinated because each of these persons will eventually enter either the *D* compartment or the *R*_*U*_ compartment, at which point they will have immunity. In the model, the effects of SARS-CoV-2 variants are captured by a time-dependent dimensionless multiplier *Y*(*t*) of the rate constant β. This rate constant, which appears in Appendix Equations 1–4, 18–22, and 24, determines the rate of disease transmission within the subpopulation unprotected by social-distancing behaviors when *Y*(*t*) = 1. We take *Y*(*t*) = 1 for times *t* < θ_1_, i.e., for the initial period of the COVID-19 pandemic in the US that we take to have started on January 21, 2020. We take *Y*(*t*) to have the form of a step function with distinct values greater than 1 for periods starting at *t* = θ_1_ and θ_*k*31_ > θ_*k*_ for *k* = 1, …, *m*. Thus, the model allows for *m* distinct periods of variant strain dominance delimited by a set of start times θ = {θ_1_, …, θ_*m*_}. We considered *m* = 2. We assume that variants differ only in transmissibility.

**Appendix Figure 2.**
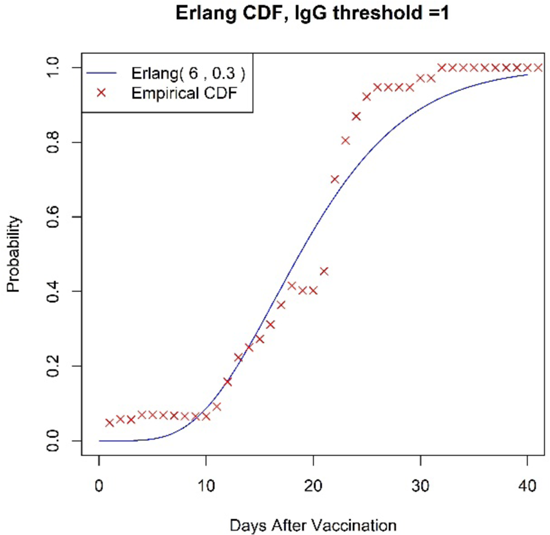
Comparison of an Erlang cumulative distribution function with shape parameter *n*_*V*_ = 6 and rate parameter *k*_*V*_ = 0.3 d^-1^ and the empirical cumulative distribution of waiting times (*t*_*V*_ values) observed in the longitudinal study of Korodi et al. [4]. The waiting time *t*^°^_*V*_ is the time between vaccination of a previously uninfected person and detection of vaccine-induced SARS-CoV-2-specific IgG antibodies.

**Appendix Figure 3.**
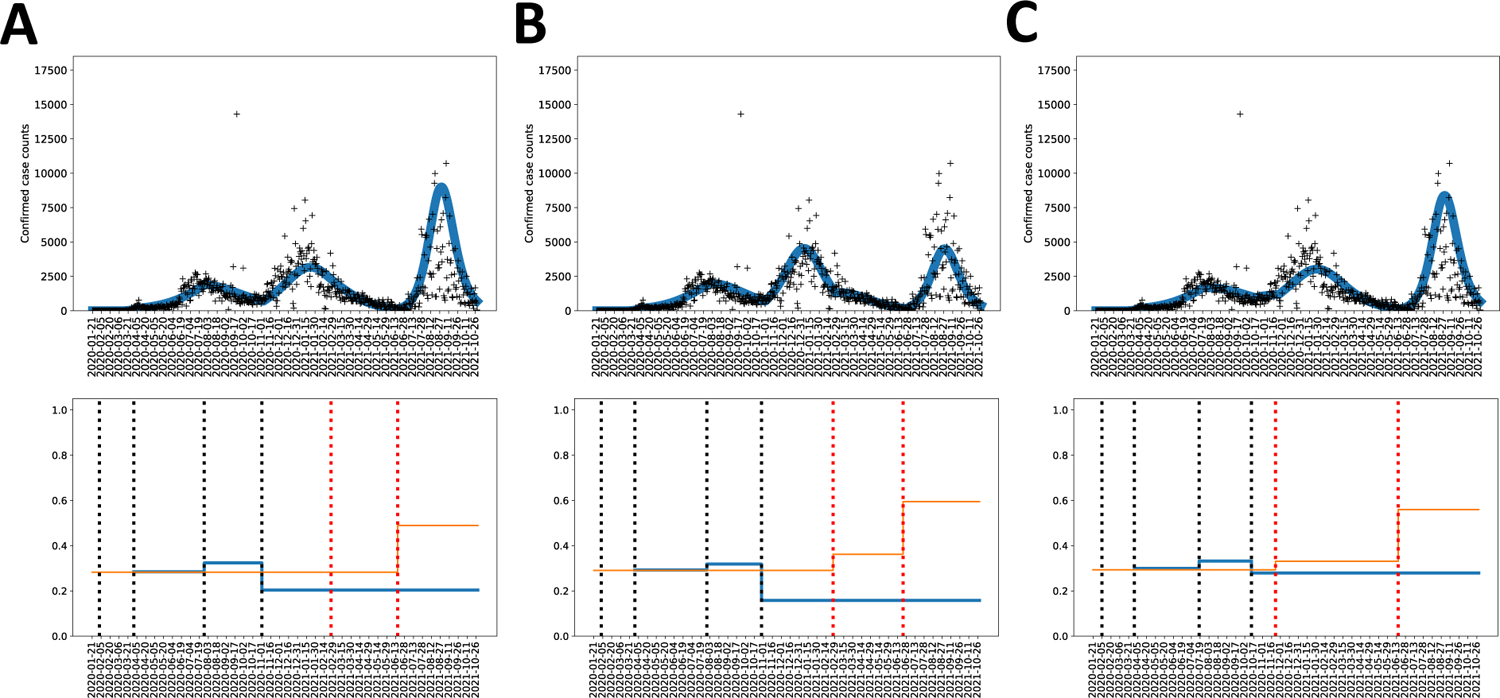
Example of a parsimony check with criteria that are violated for the MSA surrounding Houston. (A) Alpha and Delta surges (identified by sequencing data) are not explained, at least in part, by increased transmissibility, while all other criteria are satisfied. Each broken vertical black line from left to right indicates the date of onset of a social-distancing stage (i.e., with n = 3 for this illustration). Each broken vertical red line from left to right indicates the takeoff date of a variant (namely, Alpha or Delta). (B) Not all social-distancing setpoint parameter values are feasible, while all other criteria are satisfied. (C) Social-distancing changes proximal to an Alpha or Delta surge (if any) precede an increase in transmissibility, while all other criteria are satisfied. In panel A, we see that the orange segment between the two broken vertical red lines, which denotes the relative transmissibility of Alpha, is no larger than the orange segment to its left, which denotes the relative transmissibility of ancestral strains of SARS-CoV-2. In panel B, the rightmost blue segment, which denotes the social-distancing setpoint parameter value for the final social-distancing stage, has a value of roughly 0.16, which is deemed infeasible by our criterion. Finally, in panel C, the rightmost broken vertical black line is proximal to the leftmost broken vertical red line, which corresponds to the takeoff time of the Alpha surge, and the social-distancing setpoint parameter value changes prior to the Alpha surge. Note that all three panels show MLE curves, each of which constitutes an acceptable fit to the data.

## Notes

### Competing Interest Statement

The authors have declared no competing interest.

### Funding Statement

This work was supported by the LDRD program at Los Alamos National Laboratory and a grant from the National Institute of General Medical Sciences of the National Institutes of Health.

### Author Declarations

The study involves only openly available human data, which can be obtained from the GitHub repository maintained by The New York Times collecting Coronavirus (Covid-19) data in the United States (https://github.com/nytimes/covid-19-data), the Covid Act Now database collecting information about completed COVID-19 vaccinations (https://covidactnow.org/), and the COVID-19 vaccine tracker of the Democrat and Chronicle newspaper (https://data.democratandchronicle.com/covid-19-vaccine-tracker).

### Summary of Updates

Table 4 was updated. Figures 2-5 were modified for greater legibility. Various minor revisions

